# The Danish Lymphoid Cancer Research (DALY-CARE) data resource: the basis for developing data-driven hematology

**DOI:** 10.1101/2024.04.11.24305663

**Authors:** Christian Brieghel, Mikkel Werling, Casper Møller Frederiksen, Mehdi Parviz, Caspar da Cunha-Bang, Tereza Faitova, Rebecca Svanberg Teglgaard, Noomi Vainer, Thomas Lacoppidan, Emelie Rotbain, Rudi Agius, Carsten Utoft Niemann

## Abstract

Lymphoid-lineage cancers (LC: lymphoma, chronic lymphocytic leukemia, multiple myeloma, and their precursors) share many epidemiological and clinical features. To develop data-driven hematology, we gathered electronic health data and created open-source data processing pipelines to create a comprehensive data resource for Danish LC Research (DALY-CARE) approved for epidemiological, molecular, and data-driven research. We included all Danish adults registered with LC diagnoses since 2002 (n=65,774) and combined 10 nationwide registers, electronic health records (EHR), and laboratory data on a high-powered cloud-computer to develop a secure research environment. We herein exemplify how DALY-CARE has been used to develop novel prognostic markers using biobank data, real-world evidence to evaluate the efficacy of care, and medical artificial intelligence algorithms deployed directly into EHR systems. The DALY-CARE data resource allows for development of both near real-time decision-support tools and extrapolation of clinical trial results to clinical practice, thereby improving care for patients with LC.

## Background & Summary

Lymphoma, chronic lymphocytic leukemia (CLL), plasma cell dyscrasia (PCD) and their precursors are a highly heterogenous group of hematological malignancies in the blood, bone marrow and/or lymphoid tissues^1^. They span from precursor states such as monoclonal B-cell lymphocytosis (MBL) and monoclonal gammopathy of uncertain significance (MGUS), over indolent B-cell lymphomas (BCL), smoldering multiple myeloma (MM) and CLL, to aggressive high-grade BCL and PCD such as amyloid light-chain amyloidosis. Lymphoid-lineage cancers (LC) accounts for approximately 5% of all newly diagnosed cancers and more than 4% of all cancer deaths worldwide, whereas precursors such as MGUS and MBL may be identified in 5% to 12% of healthy, older individuals^2,3^.

Despite the pathological, biochemical, molecular, and clinical heterogeneity of LC, they share several common features. First, all LC derive from or are directly caused by monoclonal lymphoid-lineage expansion in blood, bone marrow and/or lymphoid tissues. Second, they are diagnosed by either histological tissue staining, flow cytometry, and/or molecular analyses, and the diagnostic workup typically includes biochemical blood tests, a bone marrow examination, and computer tomography (CT) or positron emission tomography (PET)/CT imaging. Third, different LC share common epidemiology such as older age at diagnosis and male predominance, whereas socioeconomic status and lifestyle exposures are generally not correlated with risk of developing LC. Fourth, LC develops due to dysregulation of immune function, stimulation and signaling in conjunction with accumulation of driver mutations^4^. Lastly, most patients with LC precursor states, CLL, indolent lymphoma or smoldering MM do not benefit from pre-emptive treatment and are therefore followed by a watch-and-wait strategy. If or when treatment is required, it usually includes corticosteroids, chemotherapy or immunotherapy in combination, while targeted therapies are rapidly substituting or adding to chemoimmunotherapy combinations for most LC^5^. From an epidemiological, molecular, and data-driven perspective, the commonalities between LC offer a unique opportunity for identifying common features across the different LC. This opens new possibilities regarding modelling disease outcomes using meta-learning and federated learning along with optimized transfer of molecular and genetic findings between the different LC. Achieving state-of-the-art accuracy of predictive models will likely require the joint modelling of diseases i.e., including training data from patients with lymphoma and multiple myeloma may improve predictive performance for patients with CLL.

To facilitate a shared data infrastructure to study LC, we developed the Danish LC Research (DALY-CARE) data resource allowing for clinical epidemiology, day-to-day monitoring of clinical outcomes, omics and functional analyses of available biobank samples^6^, and data-driven medical artificial intelligence (mAI) research^7^. Specifically, DALY-CARE has been approved by the Danish National Ethics Committee to collect 1) biobank samples and 2) electronic health data (EHD) retrospectively and prospectively for all Danish adult residents diagnosed with LC. DALY-CARE is approved for the study of the integrated clinicopathological profile of LC based on medical history, clinical and paraclinical (i.e. biochemical, cellular, microbiological, pathological, radiology, molecular, and genetic) routine data, while molecular omics (e.g. chip-array techniques, genomics, transcriptomics, proteomics, and microbiomics) and functional analyses (e.g. immune assays, phospho-flow cytometry, in vitro drug screening and mouse xenograft models) of adjoined biobank samples are covered by the protocol for correlations with the course of treatment and clinical outcome based on clinical epidemiology and data-driven mAI approaches.

The main driver in the recent successes of AI for image analysis and natural language processing outside of medicine is the critical mass of researchers working on the same benchmarks competitively and collaboratively. This ensures a continuous improvement of state-of-the-art performance on the given datasets^8^. By contrast, access to most medical data is heavily restricted. For instance, the performance of prognostic models in CLL has plateaued over the last 20 years likely due to lack of (1) multiple data modalities, (2) time-series modelling, and (3) a diverse set of researchers with access to the same benchmarks^9^. In turn, most mAI is developed on outcomes that do not have the highest clinical impact - but instead on the restricted data sets that are easily and publicly accessible to data scientists (e.g. the MIMIC III and IV datasets)^9,10^. With the DALY-CARE resource, we thus aim to address the aforementioned issues of lack of multimodality, common access, and clinically valid outcomes. Within DALY-CARE, we will create benchmark datasets for outcomes with high clinical impact (e.g. death, treatment response, cardiac events, adverse events and infections) and provide access to the multimodal data necessary to model these. We expect this pipeline to accelerate data-driven research for hematological malignancies and the subsequent development of state-of-the-art decision support tools. The DALY-CARE data resource provides the setup for automatized near real-time capture and monitoring of different outcomes upon changes in clinical care as well as upon deploying decision support tools, whether developed based on the DALY-CARE cohort or based on external datasets.

In this paper, we provide an overview of the DALY-CARE data resource, its infrastructure, and data formats. We exemplify the potential of the data resource by providing examples of published studies and algorithms directly based on the DALY-CARE data resource.

## Methods

### Study population

We included all Danish adult residents registered with a LC diagnosis since 1 January 2002 using International Classification of Diseases version 10 (ICD10; i.e. C81.x-C90.x, C91.1-C91.9, C95.1, C95.7, C95.9, D47.2, D47.9B, and E85.8A) and Systematized Nomenclature of Medicine (SNOMED) codes, which could be mapped to ICD10 codes for LCs (Supplemental Table S1)^11^. Patients were identified from three data sources: 1) the Danish Clinical Quality Program – National Clinical Registries (RKKP), 2) the Danish Health Data Authority (SDS), and 3) the EPIC®-based EHR system in eastern Denmark (*Sundhedsplatformen* [SP]). Nearly all Danish patients with LC are referred to one of eight Danish hematology departments, which are placed in five Danish regions (Supplementary Information: Hematological centers and regional assignment to patients), and the nationwide coverage for malignant LC diagnoses within RKKP has previously been estimated to 99%^12–14^. The cutoff date for follow-up and inclusion of data and newly diagnosed patients with LC was 15 Nov 2023.

### Data sources

EHD were primarily gathered from existing data sources as summarized in Table 1. From the RKKP registers, these include the Danish National Lymphoma Registry (LYFO)^15^ since 2005, the Danish National Multiple Myeloma Database (DaMyDa)^14^ since 2005, and the Danish National Chronic Lymphocytic Leukemia Registry (DCLLR)^12^ since 2008. Exhaustive lists of variables in the RKKP registers are described elsewhere (Supplemental Table S2)^16^. From SDS starting from 2002, existing data sources included the Register of Pharmaceutical Sales covering prescription drug data (LSR)^17^, the National Hospital Medication Register covering in-hospital medication (SMR, coverage since 2004)^18^, the Danish National Pathology Register (PATOBANK) including pathology notes (free text) and SNOMED codes^19^, the Clinical Laboratory Information System Database with routine laboratory results (LABKA)^20^, the Danish Register of Causes of Death (DAR)^21^, the Danish National Patient Registry (LPR) versions 1 and 3 with diagnosis and procedure codes^22,23^, and the Danish Cancer Registry (DCR)^24^ (Supplemental Table S2).

We retrieved EHD from 14 modules in the EPIC® (or SP) EHR system of eastern Denmark including 1) administered medicine, 2) admissions (ADT), 3) active in-hospital diagnoses, 4) all test results, 5) hematology/oncology treatment plans, 6) microbiology charts, 7) transfers between departments, 8) intensive care unit admissions, 9) medical notes (as free text), 10) prescribed medicine, 11) out- and in-patient visits and diagnoses, 12) anthropometrics, 13) a social history including smoking and alcohol consumption, and 13) vital signs (Supplemental Table S3; Supplementary Information: Go-live dates).

Additionally, we gathered EHD indirectly from laboratory systems at our institution. These included cleaned versions of the Danish Microbiology Database (MiBa)^25^ and LABKA retrieved through the Personalised Medicine of Infectious Complication in Immune Deficiency (PERSIMUNE) covering the Capital Region of Denmark (Table 1)^26^. In addition, we added summary reports and results directly from the laboratory systems for routine flow cytometry analyses^27^, immunoglobulin heavy-chain variables gene (IGHV) analyses including stereotypic subset designation^28^, targeted next-generation sequencing (tNGS)^29^, and fluorescence in situ hybridization (FISH) reports. Manually curated datasets collected through EHR reviews include information on patients receiving second line CLL treatment, patients with CLL treated with ibrutinib, patients with MM treated with daratumumab, and patients with CLL developing Richter’s transformation from previously published cohorts^30–33^.

Finally, the DALY-CARE protocol specifically allows for functional and molecular analyses including omics of biobank samples from individuals in the cohort. For this purpose, we gathered available biobank information from four large Danish biobanks, namely the CLL biobank, the Copenhagen Hospital Biobank, the Danish Cancer Biobank, and PERSIMUNE biobank (Table 1)^34^. Imputed genotypes from peripheral blood at time of first hospital blood-workup are available in 9,320 individuals (see Supplementary Information: Future data perspectives)^35^.

### Ethical approvals

The DALY-CARE protocol has been approved by the Danish Health Data Authority and National Ethics Committee (approvals P-2020-561 and 1804410, respectively). According to Danish legislation, the collection of electronic health register data is mandatory, and data were collected for research purposes in accordance with the approved protocol (see Supplemental Appendix). The Danish National Ethics Committee granted an exemption for patients to provide informed consent in order to share electronic health data. The exemption was based on the potential high impact for the patient group in question, thus considered to outweigh the issues raised by an exemption. This exemption was also extended to allow analyses of biobank samples including extensive molecular analyses for the retrospective part of the cohort, while for such prospective sampling, written informed consent was provided by patients to collect and analyze biobank samples and electronic health data.

## Data Records

### Main variables

Variables in existing data sources have been described elsewhere (Supplemental Table S2) ^12,14,15,17–21,23,25^. In short, baseline demographic, prognostic, common biochemical, and molecular data, as well as clinical data on treatment, response, and survival are assembled from the RKKP registers, which are kept as wide-format datasets and cleaned through protocols available in the DALY-CARE data resource (Supplementary Information: Software). The main variables use encoding according to the Danish Medical Classification System^a^ (SKS)^36^. In brief, SKS encoding covers diagnoses, pathology, biochemistry, locations as well as surgical, radiological and treatment procedures using ICD10, SNOMED for pathology, nomenclature property units (NPU) for laboratory, health care provider location (SHAK), and anatomical therapeutic chemical (ATC) codes for medicine (Supplementary Information: Codes and formats). These codes facilitate clear definitions and easy mapping of data when linking datasets in DALY-CARE (Supplemental Table S4-S5). All diagnoses, medicine, and biochemistry across all available datasets have been gathered into independent aggregated views.

We underscore that data do not include any sensitive information such as names, addresses, religion, ethnicity, political views, financial data, or sexual orientation.

### Database infrastructure

The DALY-CARE data resource is based on open source scalable PostgreSQL located inside a secure ISO 27001 certified private cloud on a high-performance supercomputer at the Danish National Genome Center (<u>NGC</u>) research infrastructure. The server is accessed via a Federal Information Processing Standard (FIPS) complaint virtual desktop interface (VDI) that provides full separation between users. Built specifically with data security and privacy in mind, the platform uses 2-factor authentication (2FA) and layered security approach. The platform is on-premises and does not use any components that are hosted outside the infrastructure, which ensures data security and compliance with the statutory acts^37^.

All data are pseudonymized based on a single patient ID linked to the Danish Civil Registration System’s CPR number^38^. This allows for unique identification of all patients and enables easy data linkage across all datasets while protecting individual patients’ data privacy (Figure 1a). All raw data are available directly in the DALY-CARE database. Newly retrieved data is quality assessed and previous data cuts are logged to ensure the reproducibility of all studies.

**Figure 1.**
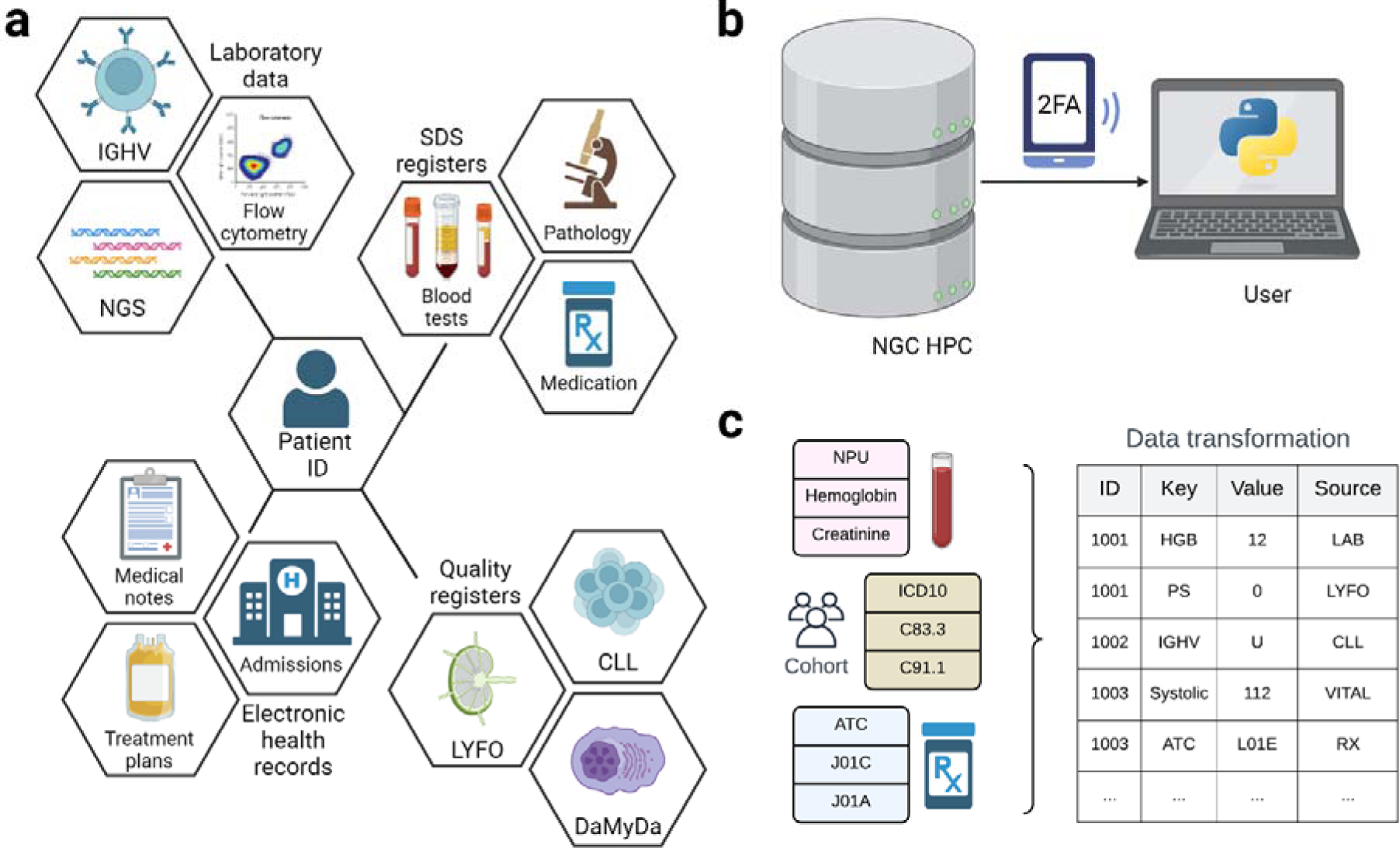
Schematic overview of the Danish Lymphoid Cancer Research (DALY-CARE) data resource. Data from various sources including Danish nationwide registers, hematological quality registers, electronic health record data, and hematological laboratories may be linked using a single pseudonymized patient ID (a). Hosted on the Danish National Genome Center high-performance computer (HPC) cloud, all data are placed in a PostgreSQL database accessed by 2-factor authentication (2FA) and loaded via R or Python software (b). Database queries and pipelines to load and transform data based on encoded data (e.g. patient ID, international classification of disease version 10 [ICD10], nomenclature property units [NPU], and anatomical therapeutic chemical [ATC]) facilitate easy data transformation commonly used for survival analyses and data-driven research (c).

The DALY-CARE server is divided into two databases with three schemas each:

1. *import*

a. *public* contains raw data retrieved from RKKP, SDS, EHR, and PERSIMUNE.
b. *laboratory* contains data from hematological laboratories including flow cytometry, FISH, IGHV, and NGS data.
c. *lookup_tables* contain tables to map encoded data including SNOMED, SKS, ATC, ICD-10, NPU, and SHAK.
2. *core*

a. *public* contains cleaned versions of raw datasets and aggregated data from multiple data sources.
b. *curated* contains manually curated datasets obtained from manual EHR reviews and NGS data.
c. *lookup_tables* contain cleaned tables to map standard coding.

We created data processing pipelines and functions for loading and processing the data in both R and Python software (Figure 1b; Supplementary Information: Software). This includes pipelines that transform the raw data into a format ready to use by predictive models (feature generation), and scripts that define and extract clinical outcomes from the raw datasets. We grouped functions based on their specific purpose inside or general use outside the DALY-CARE server to allow for synergy in larger collaboratives of Danish epidemiologists and data scientists studying similar register data as well as for adaptation to similar international data. Importantly, the pipelines in R and Python software allow for direct database queries to quickly load data using indexed encoded variables (Supplemental Table S4)^39,40^.

### Biobank material

On top of the electronic register and EHR data, we also included laboratory data as indicated in Table 1. To facilitate large-scale omics studies in DALY-CARE, we searched four large Danish biobanks and identified 40,863 biobank samples either stored on different dates or collected from different tissues from 18,528 individuals. This included 21,431 samples among 9,636 patients with lymphoma (ICD10 codes C81.x-C89.x), 11,043 samples in 3,809 patient with CLL (ICD10 code C91.1), and 6,206 samples in 3,216 patients with PCD (ICD10 codes C90.0-C90.3, D47.2, and E85.8A). The different tissues included bone marrow (n=2,535), lymph node (n=2,624), peripheral blood (n=24,068), DNA from peripheral mononuclear cells (n=2300), viably frozen cells (n=826), and plasma (n=8,510). Most peripheral blood samples were drawn upon first hospital visit and may in most instances serve as germline (normal) samples. To create a large genetic repository in the future, genotyping (n>12,000), whole genome sequencing (n>2,000), and proteomics (n>800) is ongoing.

## Technical validation

### Patients

We identified all adult patients with ICD10 codes or SNOMED codes that could be mapped to LC diagnoses (Supplemental Table S1 and S6). In total, we included 65,744 patients with a LC of whom 35,399 (53.8%) had more than one diagnosis: 15,838 (24.1%), 10,420 (15.8%), 5315 (8.1%), and 3826 (5.8%) patients had 2, 3, 4, and >4 different LC diagnoses, respectively. Most patients with multiple LC diagnoses were initially diagnosed with unspecified LC (e.g. C81.9, C82.9, C85.5, C85.9, and C91.9) followed by specified subclassification, whereas fewer patients had multiple diseases, progressed from precursor states to overt LC or transformed to more aggressive disease (Figure 2; Supplementary Information: Detailed patient information). The 20 most common LC diagnoses could be attributed to 62,946 patients (95.7%) and are summarized in Table 2.

**Figure 2.**
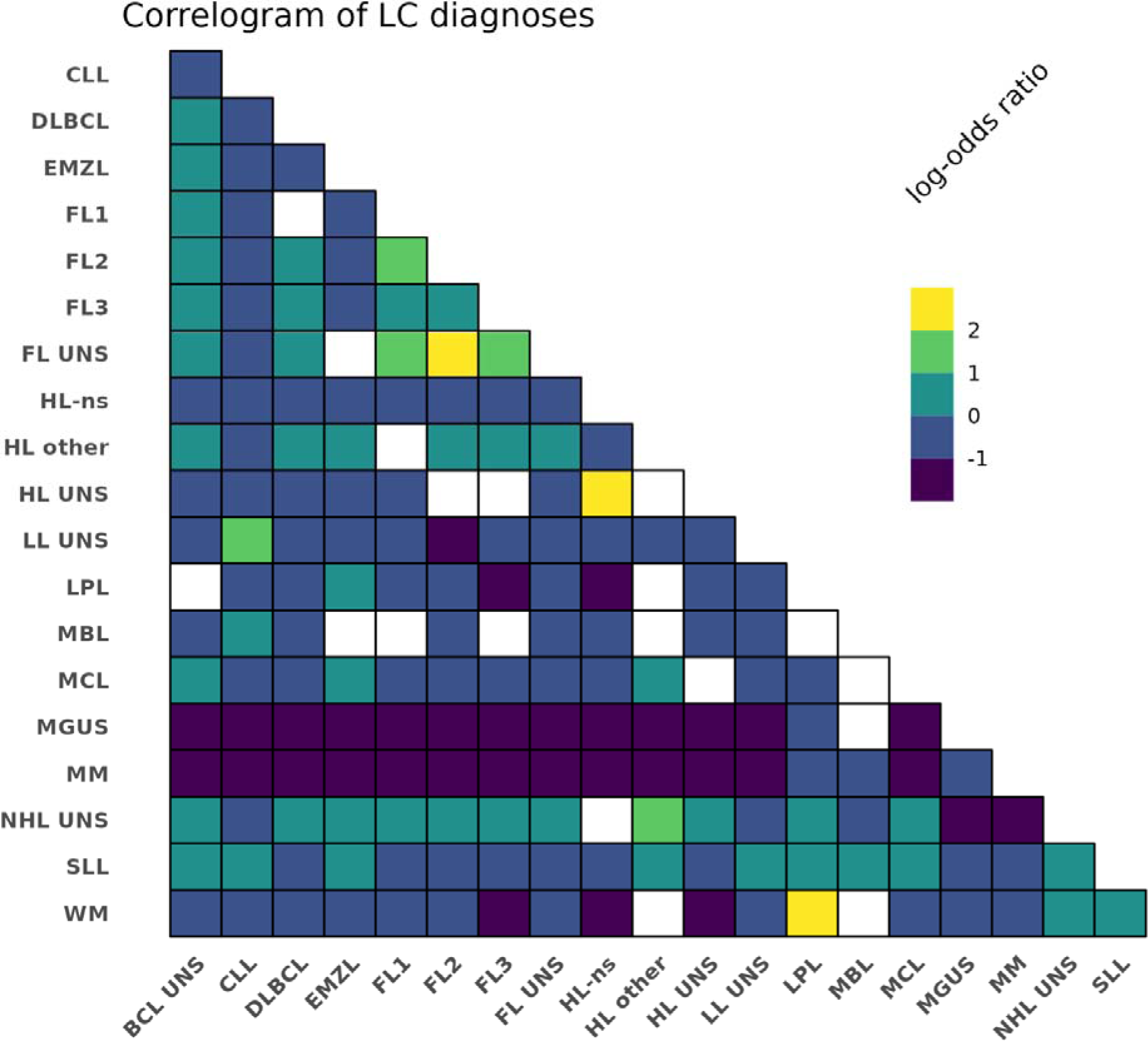
Correlations between lymphoid-lineage cancer (LC) ICD10 diagnoses shown as pairwise Fisher’s exact tests. The log odds ratio is indicated when the false detection rate (FDR) was above 0.1. ICD10 pairs with an FDR ≥0.1 are indicated in white. B cell lymphoma, BCL; chronic lymphocytic leukemia, CLL; diffuse large B cell lymphoma, DLBCL; extranodal marginal zone lymphoma, EMZL; follicular lymphoma, FL; Hodgkin lymphoma, HL; lymphoid leukemia, LL; lymphoplasmacytic lymphoma, LPL; mantle cell lymphoma, MCL; monoclonal gammopathy of uncertain significance, MGUS; multiple myeloma, MM; monoclonal B-cell lymphocytosis, MBL; non-Hodgkin lymphoma, NHL; nodular sclerosis, ns; not otherwise specified, NOS; small lymphocytic lymphoma, SLL; unspecified, UNS; Waldenström macroglobulinemia, WM.

At time of first LC diagnosis, the median age was 70.3 years (interquartile range [IQR] 60.7;77.9) and 56.1% were male (Table 3). From the cause of death register, RKKP and EHR data, we compiled dates of last follow-up (31 Dec 2020, 15 Nov 2023, and 3 Sep 2023, for each source respectively) or death to calculate time from first LC diagnosis to death or end of follow-up. In 1763 (2.7%) patients the last date of follow-up antedated the first diagnosis due to registration lag time. As a result, Kaplan-Meier days and survival status are readily available in the DALY-CARE data resource for survival analyses. The median follow-up time was 8.5 years (IQR, 4.9;13.4). For the 10 most common LCs, the 5-year unadjusted OS ranged from 50.0% in MM to 74.8% in FL (Figure 3; Supplemental Table S7). A detailed description of the patients and baseline characteristics is available in the Supplementary Information (please see Detailed patient information). We next used clinical baseline characteristics including polypharmacy defined by prescriptions in the year prior to first LC diagnosis (Supplemental Table S8) to perform multivariable Cox regression analyses in LC subtypes with available information on disease-specific international prognostic indices (IPI; Supplemental Table S9). Adjusted for age, sex, IPI, Charlson comorbidity index (CCI) score (2 vs >2), and polypharmacy (<5 vs ≥ 5 drugs), we demonstrated an independent association with shorter OS for disease-specific IPI, CCI score, and polypharmacy (Supplemental Figure S3)^41,42^. However, polypharmacy intervals (0-3, 4-6, 7-9, 10-12, and >12 different ATC codes) further demonstrated a notable dose-response effect on OS, which likely underscores an improved ability of prescriptions to explain the survival impact of comorbidity better than ICD10 codes obtained from hospital admissions used to calculate CCI scores (Figure 4).

**Figure 3.**
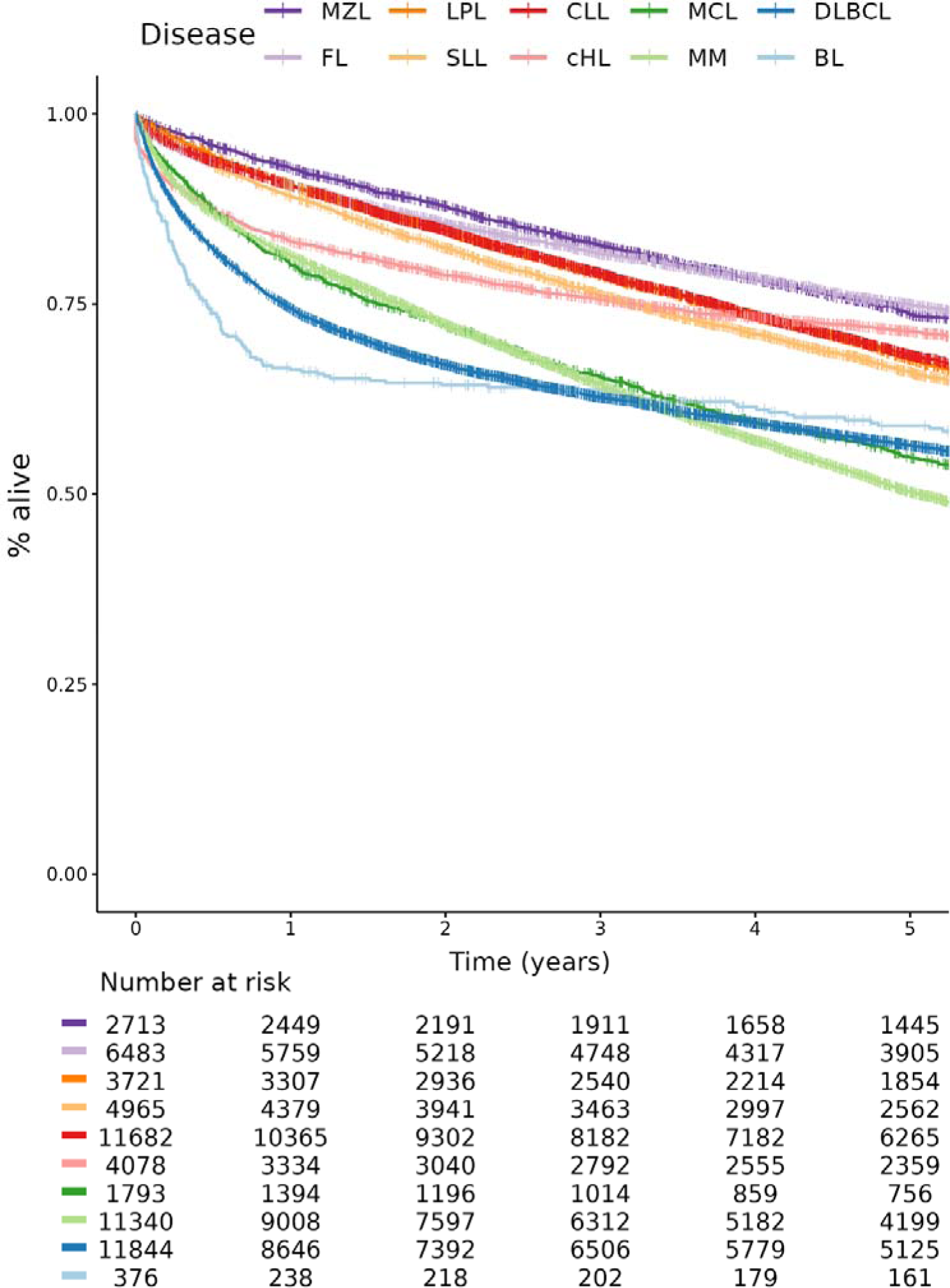
Unadjusted overall survival in 10 LC diagnoses. Burkitt lymphoma, BL; classical Hodgkin lymphoma, cHL; chronic lymphocytic leukemia, CLL; diffuse large B cell lymphoma, DLBCL; follicular lymphoma, FL; lymphoplasmacytic lymphoma, LPL; mantle cell lymphoma, MCL; multiple myeloma, MM; marginal zone lymphoma, MZL; not otherwise specified, NOS; small lymphocytic lymphoma, SLL.

**Figure 4.**
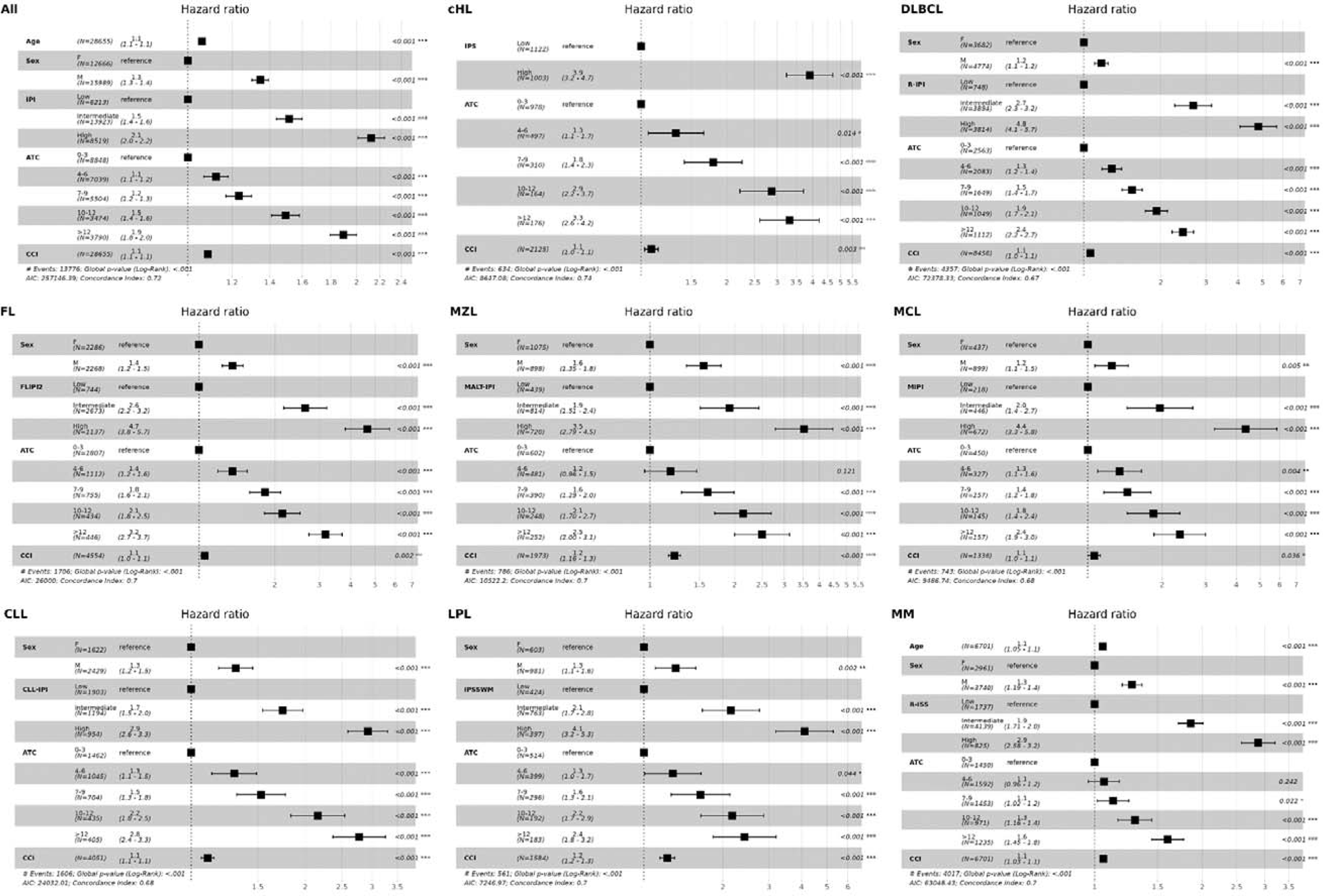
Multivariable analyses on age, sex, disease-specific international prognostic index (IPI), the number of anatomical therapeutic chemical (ATC) codes from prescription in the year prior to diagnosis, and Charlson comorbidity index (CCI) score in all disease, classical Hodgkin lymphoma (cHL), diffuse large B cell lymphoma (DLBCL), follicular lymphoma (FL), marginal zone lymphoma (MZL), mantle cell lymphoma (MCL), chronic lymphocytic leukemia (CLL), lymphoplasmacytic lymphoma (LPL), and multiple myeloma (MM). In the pooled analysis of all disease, CLL-IPI high and very high risk grouped into high IPI and revised international staging system (R-ISS) I, II, and III were labeled low, intermediate, and high risk, respectively.

### Data coverage

To avoid bias when linking different datasets, we provide an overview of coverage over time for data from RKKP, SDS, EHR, and PERSIMUNE datasets (Figure 5 and Supplementary Figure S4). While all data are population-based, data collection was centered around eastern Denmark and our institution in specific. We thus calculated the prevalence of CLL, DLBCL, and MM in the data available in DALY-CARE to assess regional differences, which would likely represent differences in data coverage (Figure 6). In addition to the spatial biases in the data, Figure 5 shows a clear temporal bias. Generally, more recent data has better coverage than older data: 13 out of 54 data sources plotted date back to 2002, while all 54 data sources have data available for 2019. Furthermore, difference in regional coverage over time reveal better coverage in the Capital Region for LABKA data sources (Supplemental Figures S5-S9 panels F1 and H1).

**Figure 5.**
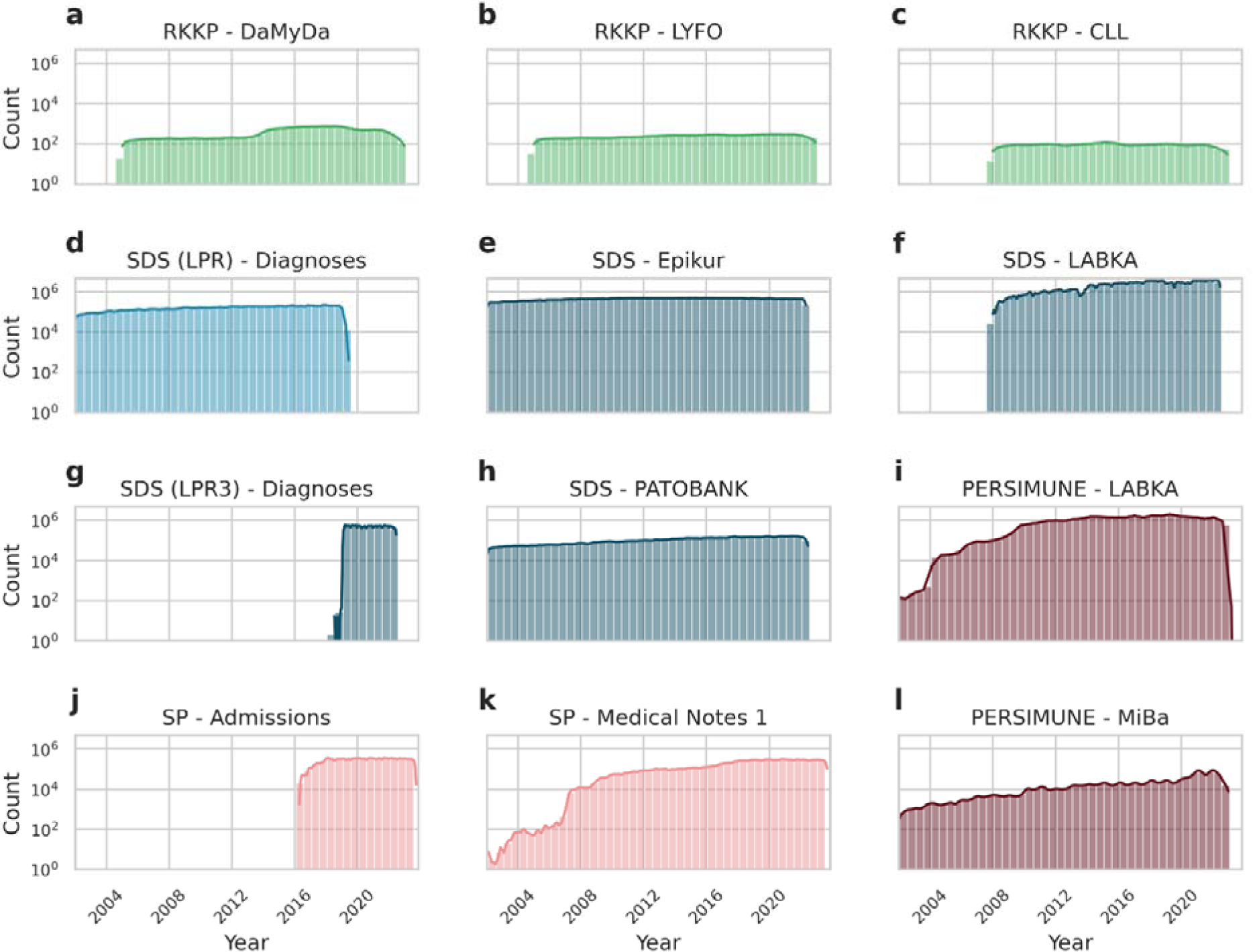
Overview of time coverage in 12 key datasets in the DALY-CARE data resource. RKKP hematological quality registers are wide format datasets, where data are typically entered upon diagnosis/registration, upon treatment, relapse, and follow-up (a-c). SDS LPR register was replaced by LPR3 in Feb 2019 (d, g). Epikur (prescriptions), LABKA (blood tests), and PATOBANK (pathology requisitions) were independent of this update (e, f, h). We underscore that similar data from different data sources may have different time coverage (f, i). The electronic health record system of eastern Denmark (SP) went live in Mar 2016 (j). Even so, medical notes antedating go-live dates from the previous EHR system were imported and are available as historic notes (k). A steadily increasing number of antimicrobial analyses was observed (l). Each panel shows the time coverage of a single dataset (a-l). Note that the y-axis shows the number of observations in three-month intervals on a log-scale. Lines above bins represent kernel density estimates of the counts.

**Figure 6.**
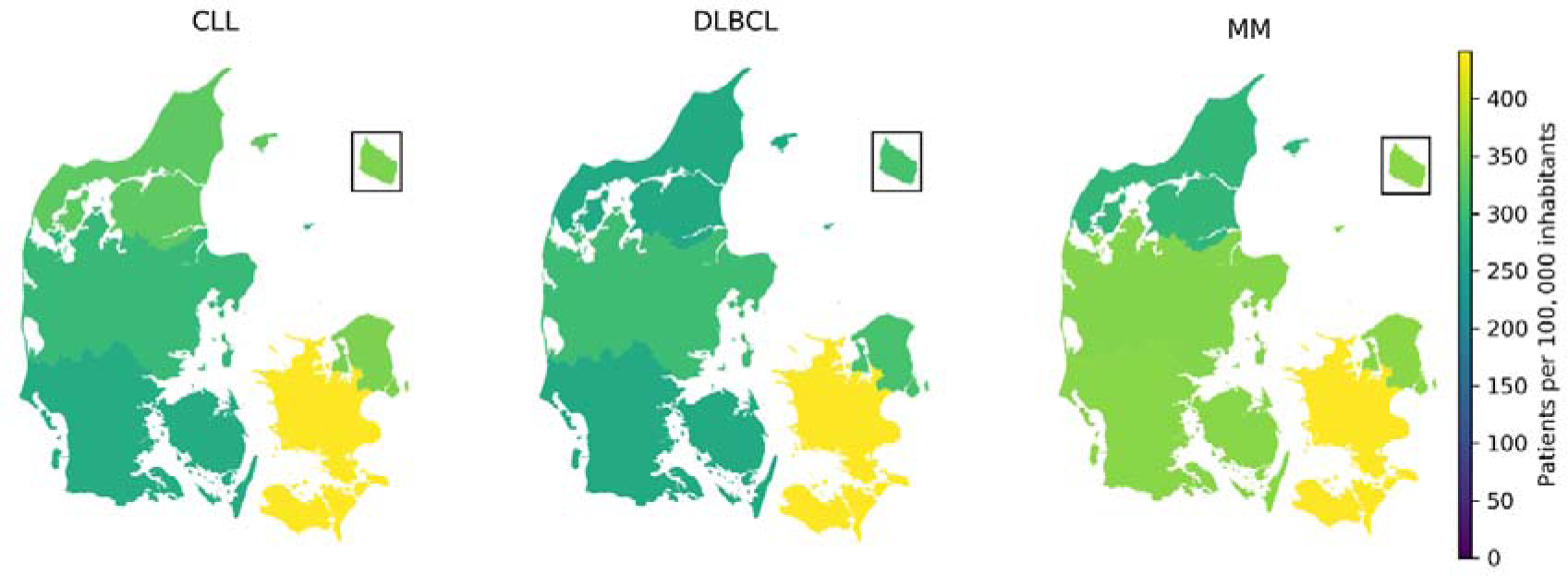
Number of patients per 100,000 residents in a) chronic lymphocytic leukemia (CLL), b) diffuse large B-cell lymphoma (DLBCL), and c) multiple myeloma (MM). Numbers included all patients in the DALY-CARE data resource regardless vital status.

### Previous data usage

Due to the variety of data sources and data modalities, the DALY-CARE data resource may facilitate a diverse set of research projects. Here we highlight some of the unique possibilities of DALY-CARE by giving examples of previously published work including use of biobank data to find novel prognostic markers, investigation of real-world evidence of different prognostic factors influencing the efficacy of health care, and development and deployment of mAI algorithms. To avoid extreme self-citing, publications have been summarized in Table 4.^43^

Using biobank data, we have validated and identified molecular risk factors associated with shorter time to first treatment and disease progression. In collaboration with the <u>European Research Initiative on CLL (ERIC)</u>, we have contributed to demonstrating adverse outcome for certain stereotypic B-cell receptor (BcR) subsets and identifying BcR subsets in approximately 40% of patients with CLL. In other ERIC collaborations, recurrent gene mutations influence time to first treatment differently in patients with mutated and unmutated IGHV status. Additionally, we have shown that recurrent gene mutations in CLL may be assigned to key signaling pathways to improve prognostication of time to first treatment. We have further demonstrated that low variant allele frequency *TP53* gene mutations do not impact overall survival time from CLL diagnosis per se, but instead have high clinical impact from time of CLL treatment, whereas patients with multi-hit *TP53* aberrations being treated with ibrutinib demonstrate shorter progression-free survival. By accessing large Danish biobanks, we have further been able to detect CLL clones in samples decades before diagnosis of CLL. In two other studies, we investigated the microbiome of CLL patients, and used fresh blood samples to demonstrate immune response improvement in clinical trial patients treated with targeted therapy. In brief, these studies highlight the potential of using multimodal approaches to achieve personalized medicine for patients based on for instance gene mutations, immune phenotype, and microbiome analyses.

With large cohorts of patients with LC included in DALY-CARE, the data resource is highly suited for real-world evidence epidemiological studies. By describing the Danish CLL register and validating the international prognostic index for CLL (CLL-IPI) in the Danish CLL population and for Binet A stage patients, we have identified variables that may supplement and improve the predictive performance of CLL-IPI. These variables include driver mutations in cell signaling pathways, eosinophil counts, hypogammaglobulinemia, comorbidity scores (CLL-CI), type 2 diabetes, and an index to identify patients without need of treatment (CLL-WONT). For CLL-WONT, we subsequently showed that it was both feasible and safe to end specialized follow-up for patients with lower risk CLL. DALY-CARE was also used to gather data from a wide range of treatment cohorts to describe real-world outcomes upon first and second line chemoimmunotherapy and ibrutinib in CLL as well as upon daratumumab in MM. In other studies, we defined and assessed the risk of clinically relevant outcomes such as second primary malignancies in CLL associated with exposure to chemoimmunotherapy, Richter’s transformation associated with CLL high risk features, atrial fibrillation upon ibrutinib, and blood stream infections in CLL and MM. Such standardized epidemiological definitions of adverse outcomes and events are available in DALY-CARE. Prior to LC diagnosis, we have also been able to demonstrate a decade long increased prescription of antimicrobials, and lymphocyte slopes up until CLL diagnosis further informing disease trajectories. Finally, during the corona virus disease pandemic, we could quickly assess and monitor mortality in a wide range of LCs, and we further demonstrated poor vaccine responses in CLL and better clinical outcomes during the omicron era. This underscores how frequently updated data allows for near real-time monitoring of large-scale real-world clinical outcomes.

We also employed DALY-CARE to develop data-driven studies using mAI. For instance, we showed that adding paraclinical information to CLL-IPI variables improved predictive performance, whereas recurrent gene mutation information did not. This highlights the potential within DALY-CARE to estimate which classes of variables and modelling approaches are most informative for a given outcome. Notably, we have developed and deployed the data driven model CLL-TIM for predicting whether patients with newly diagnosed CLL will have an infection or will be treated within 2 years. This model was implemented into the ongoing PreVent-ACaLL clinical trial (<u>NCT03868722</u>) and the EHR system of eastern Denmark, and we have shared recommendations and considerations for easing transition from development to deployment for future mAI algorithms^44^. In conclusion, these studies highlight that DALY-CARE facilitates transformation of detailed EHD and biobank-based analyses into actionable models and prognostic indices with direct impact on daily clinical practice.

## Discussion

The DALY-CARE data resource is a collection of national health registries, EHRs, routine and special laboratory data as well as analyses from adjoined biobanks. Including more than 65,000 patients with LC, this data resource was gathered and stored on a super-computer cloud ensuring both the data privacy and safety, storing capacity, and the processing power needed to handle sensitive data without compromising the risk of data migration^45^. Based on published studies using the DALY-CARE data resource, we provide examples of how DALY-CARE can lead to (1) finding novel prognostic markers using biobank data, (2) using real-world evidence studies to evaluate the efficacy of routine health care, and (3) deploying mAI algorithms directly into EHR systems.

The DALY-CARE cohort is truly population-based as nearly all Danish adult LC patients are referred to one of eight hematological centers and cancer registration is mandatory by law. This results in a register coverage of 99%^13^. Even so, we found large regional disparities in the available data for patients mainly owing to EHR data being available for only two out of five Danish Regions, covering half of the Danish population. The use of ICD10 codes upon admissions may also differ widely across institutions. This skew in availability and usage could limit the scope of the possible studies or produce unintentional biases in data driven algorithms if they are not accounted for. Like other EHD archives, data in DALY-CARE are affected by shifts in systems such as the transition from LPR to LPR3 (i.e. 2 Feb 2019) or implementation of EPIC® in eastern Denmark (from May 2016 through Nov 2017; Supplementary Information: Go-live dates). These time dependent changes in data and data formats are important to recognize to avoid biases that could lead data driven algorithms astray. To this end, we focused on 1) describing when and where the data are available and identified sub-cohorts according to ICD10 diagnoses, 2) standardization and quality control for measurements (i.e. making the same measures using different units commensurable), and 3) using robust clinical definitions to calculate features based on domain-knowledge such as prognostic indices.

By highlighting previously published work (Table 4), we hope that DALY-CARE will serve as an important first step towards standardizing and creating similar data resources for other research groups. This would help alleviating the problem of limited training data by enabling the use of techniques that can leverage information learned from training on other cohorts such as meta-learning^46^. Beyond the national generalization (e.g. SKS coding), we mapped 90% of our curated data from medicine, laboratory measurements and diagnosis to OMOP standardization to facilitate international collaborations. This will also serve as a critically important step for implementing state-of-the-art mAI that aggregate information across countries using federated learning.

Most research in mAI is almost exclusively focusing on developing novel methods and algorithms, whereas reports of deploying, monitoring, improving, or maintaining existing algorithms are anecdotal^47,48^. Outside of image analysis used for clinical care, mAI is rarely deployed directly into clinical patient care. The DALY-CARE data resource provides real-world clinical data and is regularly updated. Essential parts of the codebase are publicly available on <u>Github</u> and regularly updated. We believe that access to near real-time real-world clinical data will ease the transition from development to production, aiding the issue of deploying algorithms.

Medical interventions for LC patients are often implemented without scientific evidence, and many interventions will never be investigated in randomized clinical trials (RCT)^49^. DALY-CARE provides means to qualify and inform the impact of changes in health care practice that have not, cannot, and likely will never be tested sufficiently in RCTs. This is exemplified by monitoring overall survival, health care utilizations and infections for patients ending specialized follow-up or receiving immunoglobulin replacement therapy. Having access to large-scale multimodal data allows not only for monitoring of clinical impact after clinical practice changes in management and supportive care but also when introducing novel treatments tested in RCT to RCT-ineligible patients. For instance, half of all Danish patients with newly diagnosed MM did not qualify for inclusion in RCT^50^, and Danish patients on ibrutinib outside clinical trials only have a median duration to discontinuation of 3 years as compared to 7 years in clinical trials^31,50,51^.

Improvements in patient care is intrinsically tied to the ability to access and share data easily^52^. Here, we demonstrate how DALY-CARE allows researchers to identify complex patterns across different patient cohorts and subgroups on a collaborative basis. The identification of such patterns may provide new data-driven hypotheses and prognostic markers as well as inform and qualify the impact of implemented management of LC patients, while providing the basis for development of improved decision support tools for LC patient care. Providing the ability to share definitions, outcomes and ultimately data will allow for competitive modeling on outcomes that would most likely increase predictive performance of prognostic models. To translate these data-driven perspectives into benefits for patients on a national and international level, we believe it is crucial to break down the arbitrary division between primary and secondary use of EHD. That is, utilizing daily routine EHR data and national registers for the purpose of clinical epidemiology, precision medicine and to enrich omics analyses with clinical data.

In conclusion, we integrated real-world data from quality hematology registers, nationwide health registers, electronic health record data, analyses of biobank samples, and data from specialized hematological laboratories to facilitate cutting edge research in clinical epidemiology, large-scale observational studies, omics analyses, and development of decision support tools based on machine-learning algorithms. Capturing and utilizing routine EHD to evaluate changes in medical practice that are not based on evidence from randomized clinical trials can now be monitored in near real-time. DALY-CARE thus paves the way for truly data-driven hematology and provide proof-of-concept for improved data-driven health care.

## Data Availability

According to Danish legislation, the collection of electronic health register data is mandatory, and data were collected for research purposes in accordance with the approved protocol (see Supplemental Appendix). The Danish National Ethics Committee granted an exemption for patients to provide informed consent in order to share electronic health data. The exemption was based on the potential high impact for the patient group in question, thus considered to outweigh the issues raised by an exemption. This exemption was also extended to allow analyses of biobank samples including extensive molecular analyses for the retrospective part of the cohort, while for such prospective sampling, written informed consent was provided by patients to collect and analyze biobank samples and electronic health data.
Due to pseudonymized but not fully anonymized nature of data, access to the data resource will be based on a Data Usage Agreement (named Data Processor Agreement [DPA], see Supplemental Appendix 1 for template). The DPA will specify the analyses performed on a collaborative basis, these analyses should be within the approved DALY-CARE protocol (Supplemental Appendix 2). All analyses must be performed on the DALY-CARE data resource at NGC. Except for pathology notes and medical notes, dummy tables are presented in Supplemental Appendix 3.
The underlying code for this study is available on the DALY-CARE data resource and can be accessed on a collaborative basis on reasonable request to the corresponding author. Essential parts of the codebase are publicly available on Github and regularly updated.

## Usage Notes

Due to pseudonymized but not fully anonymized nature of data, access to the data resource will be based on a Data Usage Agreement (named Data Processor Agreement [DPA], see Supplemental Appendix 1 for template). The DPA will specify the analyses performed on a collaborative basis, these analyses should be within the approved DALY-CARE protocol (Supplemental Appendix 2). All analyses must be performed on the DALY-CARE data resource at NGC. Except for pathology notes and medical notes, dummy tables are presented in Supplemental Appendix 3.

## Code availability

The underlying code for this study is available on the DALY-CARE data resource and can be accessed on a collaborative basis on reasonable request to the corresponding author. Essential parts of the codebase are publicly available on <u>Github</u> and regularly updated.

## Author contributions

C.U.N. conceived the project. C.U.N., C.B., and C.M.F. collected the data. C.M.F. created the database. C.B., C.M.F, M.W., and T.L. created the database infrastructure. M.W. and T.L. performed data quality control. C.B. and M.W. wrote the draft manuscript. All authors contributed to and approved the final manuscript.

## Acknowledgements

We would like to thank dr. Peter de Nully Brown for founding the Danish National Lymphoma Registry and leading the way for Danish quality registers in hematology. We sincerely thank Anders Christian Riis-Jensen and Anton Kokholm Andersen from CØK within the Capital Region of Denmark for retrieving and providing EHR data. We further thank Professor Jens Lundgren leading PERSIMUNE for providing laboratory and microbiology data, Professor Sisse Rye Ostrowski for collaboration with the Copenhagen Hospital Biobank, dr. Ida Schjødt for kindly providing flow cytometry data from the Surface Marker Laboratory at Rigshospitalet, dr. Mette Klarskov Andersen for kindly providing cytogenetics data from the Department of Clinical Genetics, Rigshospitalet, and Lone Bredo Pedersen for performing and providing IGHV analyses from the CLL laboratory, Rigshospitalet.

## Footnotes

a Sundhedsvæsenets Klassifikations System

## Tables

Please see separate excel files.

Table 1. Overview of datasets in the DALY-CARE data resource

Table 2. Most common lymphoid-lineage cancer disorders in the DALY-CARE data resource.

Table 3. Patient characteristics.

Table 4. DALY-CARE data resource usage.

