## Supplemental Information for "The Danish Lymphoid Cancer Research (DALY-CARE) data resource: the basis for developing data-driven hematology"

### Hematological centers and regional assignment to patients

Denmark is divided into five Danish Regions (Capital [Hovedstaden; SHAK 1300-2499 + 4001], Zealand [Sjælland; SHAK 2500-3999], Southern Denmark [Syddanmark; SHAK 4200-5999 + 6008], Central Denmark [Midtjylland; SHAK 6006 + 6500-7999], and North Denmark [Nordjylland: 8000-8999]), which are politically and economically responsible for providing secondary and tertiary health care in Denmark^1,2^. The Danish Regions harbor eight different hematological centers (SHAK specialty 004); excluding the Department of Hematology at Herlev Gentofte Hospital – Herlev Hospital (SHAK 1516), which was merged with Rigshospitalet (SHAK 1301) on 14 April 2021. Using available SHAK codes^2^ (Supplemental Table S2 and S4), we assigned regional information to as many patients as possible. However, not all data sources have regional information available. For this reason, we use information available in other datasets to estimate regions when not available. This was done by assembling all the data and sorting based on the date provided in the dataset. Using available SHAK codes^2^ (Supplemental Table S2 and S4), we assigned regional information data sources by forwards- and backwards filling the regional information when no regional information was provided.

For the datasets from PERSIMUNE, data were collected for patients with hospital contact in the Capital Region. Thus, if these patients had blood workup or microbiological tests performed at hospitals outside the Capital Region, these data would also be contained in PERSIMUNE data.

### Go-live dates

Data from different hospitals were available from different time points since the hospitals had different dates of implementing the EPIC®-based EHR system (SP). These so-called go-live dates were 21 May 2016 at Copenhagen University Hospital - Herlev and Gentofte, 5 November 2016 for Copenhagen University Hospital – Rigshospitalet, and 25 November 2017 for Zealand University Hospital – Roskilde. Even so, historic EHR medical notes (free text) from the previous EHR system may antedate go-live (OPUS®). Last date of follow-up including vital status within the EHRs was provided within the ADT dataset and specified as SP_OS (Supplemental Table S3).

### Future data perspectives

On top of the data assembled in the DALY-CARE data resource and described in the main text, additional data are in process and soon expected to be included in the data resource. These data include imputed genotype data from already performed on peripheral blood samples from 11,979 patients,^3^ whole genome sequencing from approximately 2,000 patients with chronic lymphocytic leukemia (CLL), proteomic analyses in approximately 800 patients with CLL, all available diagnostic fluorescence in situ hybridization (FISH) analyses from the Capitol Region of Denmark from patients with CLL, lymphoma, and plasma cell dyscrasia (PCD), and transfusion history and radiological notes from the EPIC®-based EHR record system of eastern Denmark. Soon, we also expect descriptions from radiological examinations such as chest X-rays, CT, and PET/CT scans. Further, a protocol amendment to also include clonal myeloid neoplasms (ICD10 C92.x-C96.x) is under consideration by the Danish National Ethics Committee.

### Codes and formats

The different datasets use different standardized codes to refer to different diagnoses, procedures, and examinations. On top of anatomical therapeutic chemical (ATC) codes (SKS M codes) and ICD-10 codes (SKS D codes), SKS codes also include procedure codes for surgery (SKS K codes) as well as other procedures and examinations (SKS B, E, and U codes). Further, the nomenclature, properties, and units (NPU) codes identify distinct biochemical analyses and health care provider^[1]^ (SHAK) codes may map any Danish hospital and department as well as private health care providers (Supplemental Tables S4 and S5). Further, pathology data use SNOMED codes, and microbiology data from the PERSIMUNE use microorganism (MORG) codes^4,5^. Overall, these coding systems are aligned with data within the EHR system, however, point of care tests (e.g. arterial blood gas and electrocardiograms) and non-pharmacological therapies such as intravenous fluids are included by name (e.g. “Ringer’s Lactate 1000 ml”), but not encoded. Exhaustive lists of coding tables are available in the DALY-CARE data resource (core – *lookup_tables*). All variables within SDS, RKKP and PERSIMUNE datasets are available in Supplemental Table S2 with an online and up-to-date description of each variable from the data provider^2,6-9^, whereas a description of variables in the fourteen EHR modules is provided in Supplemental Table S3. Among laboratory datasets from our institution, IGHV analyses include information on germline ID and B-cell receptor stereotypic subsets^10,11^, FISH data contain summary text and number of counted probes, flow cytometry data contain summary text and tables of immune phenotypic subpopulations^12^, and tNGS data are provided as both summary files and sequencing outcome files (i.e. bam and vcf formats).

Whereas ATC, SNOMED, ICD10, and SKS coding systems are built logically, the NPU codes (>25,000 distinct) have been numbered historically – sometimes including multiple codes for the same biochemical measurement (e.g. creatinine) due to analyses at different centers or with different biochemistry. To alleviate this problem, we grouped NPU codes to define biochemical analyses commonly used in routine clinical practice and grouped single analyses into sets of blood tests such as complete blood counts and infection sets allowing for easy identification of disease-specific biochemical markers (Supplemental Table S5).

### Software

On top of R and Python software, analyzing the larger data files for genomic data effectively requires specialized software. Integrative Genomic Viewer (IGV; Broad Institute, Cambridge, MA, USA) and VarSeq version 2.4.2 (Golden Helix, Bozeman, MT, USA) software are available to ensure plans to compile a large repository for WGS data within the DALY-CARE database including NGS output files such as bam and vcf formats. Hosted on the Danish National Genome Center’s high-performance computer, the DALY-CARE data resource relies on validated GATK-based pipelines. Retrieving results and output files by individual users is allowed only for small image-files that may be downloaded via a separate secure file transfer protocol (SFTP) server accessed by yet another 2-factor authentication (2FA) system, which alerts data management with a copy. Further, we built the *dalycare* package, which a group of functions to load, clean and filter data and to define and generate features for specific clinical outcomes. The *dalycare* package is available in the DALY-CARE data resource and on [GitHub](https://github.com/RH-CLL-LAB/dalycare_package/tree/main/dalycare). The random_dummy_tables function was used to create dummy tables available in Appendix 3.

### Detailed patient information

In addition to the demographic information provided for the patients, we tested correlations between these common ICD10 diagnoses using pairwise Fisher’s exact tests, and while most pairs were mutually exclusive of one another, we found strong co-occurrence of WM:LPL, FL_UNS:FL1, FL_UNS:FL2, FL_UNS:FL3, FL1:FL2, HL_UNS:HL-ns, NHL_UNS:HL other, and CLL:LL_UNS (odds ratio [OR] >10; Figure 3A). In detail, CLL:SLL demonstrated a weaker but significant cooccurrence (OR 4.08), whereas CLL:DLBCL were mutually exclusive (OR 0.19) despite 594 of 11,724 CLL patients (5.1%) with potential Richter’s transformation. MBL and CLL demonstrated a weak cooccurrence (OR 1.27), whereas MM:MGUS to our surprise were mutually exclusive (OR 0.77) likely indicating that most patients with newly diagnosed MM do not progress from known and/or registered MGUS. In general, both MGUS and MM mutually excluded every common LC diagnosis (Figure 2A).

We gathered all LC ICD10 codes (i.e. C81.x-C90.x, C91.1-C91.9, C95.1, C95.7, C95.9, D47.2, D47.9B, and E85.8A) or SNOMED codes mapping to LC ICD10 codes (Supplemental Table S1) from 1 Jan 2002 in adult individuals of at least ≥18 years of age at time of diagnosis. First LC diagnosis was defined as the first occurrence of a unique ICD10 code (e.g. D47.2 or MGUS) regardless of later progression (e.g. C90.0 or MM) or transformation (e.g. C90.1 or PCL) or second LC diagnosis (e.g. C83.3 or DLBCL). To include adult patients only, we excluded ICD10 codes matching a first LC diagnosis acquired before the age of 18 years, while diagnoses obtained from the Pathology Register (PATOBANK) after 18 years of age were included regardless of whether the same diagnosis was also registered before 18 years of age (Supplemental Figure S1). Patients were followed from first LC diagnosis until death or end of follow-up, whichever came first. Date of death was extracted from the Cause of death registry (DAR), RKKP registers, and SP ADT. Date of last follow-up was assigned according to the data extraction date of either of these three registries, whichever came last. In case of discrepancies between date of death, we used a hierarchy where date of death was preferred from the DAR over RKKP registers over SP ADT (Supplemental Figure S2).

From all ICD10 diagnoses (n = 14,072,561 in total, median 31 unique diagnoses per patient [IQR 21;44]) and all prescriptions (last follow-up 1 May 2022, n=18,318,049 Rx [1172 distinct ATC codes]), the median Charlson comorbidity index (CCI) score prior to first LC was 2 (IQR, 2;3: including the hematological 2 point score attributed to the LC), and the median number of prescribed drugs the year prior to first LC in 62,766 patients diagnosed before May 2022 was 6 (IQR 3;10), respectively^13,14^. As a result, 38,501 (61.3%) patients had polypharmacy (i.e. ≥5 drugs; Table 3)^14^.

Disease entities (e.g. classical Hodgkin lymphoma [cHL] and follicular lymphoma [FL]) were grouped based on ICD10 codes (Supplemental Table S7). Baseline characteristics for CLL, SLL, MCL, LPL, MZL, FL, DLBCL, cHL, MM, and MGUS are provided in Supplemental Table S8. Except for cHL, BL, and FL, the median age at first diagnosis was 69.4 to 72.9 years. The median age for FL, BL, and cHL was lower: 66.1 years (IQR 57.1;74.3), 58.7 years (IQR 42.6;71.1), and 56.4 years (IQR 35.0;70.4), respectively. All entities had male predominance except for MZL. The median CCI score was 2 in all disease subgroups (including the hematological score of 2 points at time of LC diagnosis), whereas polypharmacy ranged 45.4% in cHL to 69.4% in MM. Next, we calculated disease-specific international prognostic indices (IPI) and scores (IPS) for patients in hematological quality registers (RKKP) with information on e.g. clinical stage (Supplemental Table S9)^15-22^.

### Data Coverage

As shown in Figure 4, the data coverage in LPR registers had abrupt cuts on 19 February 2019 when LPR transitioned to and replaced LPR3. By contrast, other SDS datasets such as LSR, PATOBANK, and LABKA had long time periods with a clear trend for increasing observations in later years. As expected, time lag registration was observed in all datasets, except for EHR datasets, for which the data feed is real-time and extracted with short notice from EPIC® upon request. Although go-live dates most often defined the earliest observations in EHR datasets, diagnoses in the Active Problems and Diagnoses dataset (Table 1) may be registered back in time (with a serious risk of introducing bias), whereas medical notes and hospital prescription data had been imported from previous EHR system, thus antedating go-live dates.

### Supplemental Figure legends

**Figure S1.** Flowchart defining the all DALY-CARE diagnoses. These diagnoses are ICD10 codes and SNOMED codes that translates into C81.x-C90.x, C91.1-C91.9, C95.1, C95.7, C95.9, D47.2, D47.9B, and E85.8A from all data tables containing ICD10 and SNOMED codes. The data table Aktive_Problemliste_Diagnoser was excluded due to risk of serious time registration bias. To include adult lymphoid-lineage cancer (LC) only, diagnoses acquired before the age of 18 years were excluded, while diagnoses obtained from the Pathology Register (PATOBANK) after 18 years of age were included regardless of whether the same diagnosis was also registered before 18 years of age.

**Figure S2.** Flowchart defining date of death or last follow-up. All data tables containing date of death were used to define date of last follow-up, whichever came last, which facilitates statistical survival analyses.

**Figure S3.** Multivariable analyses on age, sex, disease-specific international prognostic index (IPI), polypharmacy (i.e. >5 different prescribed drugs within the year up until lymphoid-lineage cancer [LC] diagnosis), and Charlson comorbidity index (CCI) score in all LC disease, Hodgkin lymphoma (HL), diffuse large B cell lymphoma (DLBCL), follicular lymphoma (FL), marginal zone lymphoma (MZL), mantle cell lymphoma (MCL), chronic lymphocytic leukemia (CLL), lymphoplasmacytic lymphoma (LPL), and multiple myeloma (MM). In the pooled analysis of all disease, CLL-IPI high and very high risk grouped into high IPI and R-ISS I-III were labeled low, intermediate, and high risk.

**Figure S4**. Overall overview of time coverage in primary datasets in the DALY-CARE data resource. Each panel shows the time coverage of a single dataset. Panels are indexed alphabetically and numerically with a character from A-O indicating the row and a number from 1-3 indicating the column. For all panels, the X-axis shows the time when observations were made, and the y-axis shows the number of observations per month on a log-scale. Three months of data are aggregated per bin. Lines drawn over bins represent kernel density estimates of the counts. To the right of each panel, the number of unique patients (N) for each dataset is shown.

**Figure S5**. Region Southern Denmark time coverage of data in primary datasets in the DALY-CARE data resource. Each panel shows the time coverage of a single dataset. Panels are indexed alphabetically and numerically with a character from A-O indicating the row and a number from 1-3 indicating the column. For all panels, the X-axis shows the time when observations were made, and the y-axis shows the number of observations per month on a log-scale. Three months of data are aggregated per bin. Lines drawn over bins represent kernel density estimates of the counts. To the right of each panel, the number of unique patients (N) for each dataset is shown.

**Figure S6**. Capital Region time coverage of data in primary datasets in the DALY-CARE data resource. Each panel shows the time coverage of a single dataset. Panels are indexed alphabetically and numerically with a character from A-O indicating the row and a number from 1-3 indicating the column. For all panels, the X-axis shows the time when observations were made, and the y-axis shows the number of observations per month on a log-scale. Three months of data are aggregated per bin. Lines drawn over bins represent kernel density estimates of the counts. To the right of each panel, the number of unique patients (N) for each dataset is shown.

**Figure S7.** Region Central Denmark time coverage of data in primary datasets in the DALY-CARE data resource. Each panel shows the time coverage of a single dataset. Panels are indexed alphabetically and numerically with a character from A-O indicating the row and a number from 1-3 indicating the column. For all panels, the X-axis shows the time when observations were made, and the y-axis shows the number of observations per month on a log-scale. Three months of data are aggregated per bin. Lines drawn over bins represent kernel density estimates of the counts. To the right of each panel, the number of unique patients (N) for each dataset is shown.

**Figure S8.** Region North Denmark time coverage of data in primary datasets in the DALY-CARE data resource. Each panel shows the time coverage of a single dataset. Panels are indexed alphabetically and numerically with a character from A-O indicating the row and a number from 1-3 indicating the column. For all panels, the X-axis shows the time when observations were made, and the y-axis shows the number of observations per month on a log-scale. Three months of data are aggregated per bin. Lines drawn over bins represent kernel density estimates of the counts. To the right of each panel, the number of unique patients (N) for each dataset is shown.

**Figure S9**. Region Zealand time coverage of data in primary datasets in the DALY-CARE data resource. Each panel shows the time coverage of a single dataset. Panels are indexed alphabetically and numerically with a character from A-O indicating the row and a number from 1-3 indicating the column. For all panels, the X-axis shows the time when observations were made, and the y-axis shows the number of observations per month on a log-scale. Three months of data are aggregated per bin. Lines drawn over bins represent kernel density estimates of the counts. To the right of each panel, the number of unique patients (N) for each dataset is shown.

1. SygeHus Afdelings Klassifikationer
