## Appendix 1 for "The Danish Lymphoid Cancer Research (DALY-CARE) data resource: the basis for developing data-driven hematology"

Case number:

**DATA PROCESSING AGREEMENT**

For the Data Processor’s processing of personal data related to the parties’ agreement of xx.xx.202x dealing with analysis on INSERT regarding the project approved by the National Committee on Health Research Ethics (NVK) - case nr.: 1804410 which entails that the Data Processor shall process personal information on behalf of the Data Controller within the Data Controller’s health science research project with the title “A Research Biobank and Clinical Database of patients with Lymphoproliferative Malignancies “(DALYCA) – formerly known as “A Research Biobank of Patients with Chronic Lymphocytic Leukemia (CLL)” (”Principal Agreement”)

Between

The Capital Region Of Denmark

Kongens Vænge 2

Cartsten Utoft Niemann

Page 1

3400 Hillerød

CVR-no: 29190623]

(”*Data Controller”*)

and

Company Name

Address

Department (if relevant)

Contact person

Company reg. number

(”*Data Processor”*)

### 1. Background to The Data Processing Agreement

1.1. This data processing agreement, including appendixes and any supplements (”Data Processing Agreement”), concerns the Data Processor’s obligation to act in accordance with the EU Parliament and Council’s regulation 2016/679 of 27 April 2016 on the protection of natural persons with regard to the processing of personal data and on the free movement of such data, and repealing Directive 95/46/EC (“General Data Protection Regulation”), as well as legislation on supplementary provisions to the regulation on the protection of natural persons with regard to the processing of personal data and on the free movement of such data (“General Data Protection Law”).
1.2. The Data Processing Agreement is an integral part of the Principal Agreement.

1.3. In case of discrepancies between the provisions in the Data Processing Agreement and any corresponding provisions in other agreements between the Data Controller and the Data Processor, including the Principal Agreement, in relation to processing of personal data, the provisions in the Data Processing Agreement take precedence. This applies regardless of anything agreed about precedence elsewhere. If, however, the Data Processor is bound by more stringent obligations in other agreements between the Data Controller and the Data Processor, including the Principal Agreement, the Data Processor must fulfil those more stringent obligations.

### 2. Data Processor’s responsibility

2.1. The Data Processor must only process personal data according to documented instructions from the Data Controller, and only to the extent that is necessary for the Data Processor to fulfil the obligations of the Principal Agreement and the Data Processing Agreement, unless processing is required under EU or Member State law to which the Data Processor is subject. In this case, the Data Processor must inform the Data Controller of that legal requirement before processing, unless that law prohibits this notification on important grounds of public interest, see General Data Protection Regulation article 28, section 3, paragraph a.

The Data Processing Agreement is part of the Data Controller’s instructions to the Data Processor. The Data Processor processes personal data on behalf of the Data Controller, and must not process personal data covered by the Data Processing Agreement for own purposes.

2.2. If the Data Processor is subject to the legislation of a third country, the Data Processor must immediately inform the Data Controller in writing if this legislation prevents the Data Processor from complying with the Data Processing Agreement and associated instructions.
2.3. The Data Processor must immediately inform the Data Controller if an instruction is, in the Data Processor’s opinion, in contravention of the General Data Protection Regulation, the General Data Protection Law or the data protection provisions of EU or Member State law (combined, “General Data Protection Legislation”)
2.4. The Data Protection Agreement does not release the Data Processor from obligations directly imposed by the General Data Protection Regulation or any other current legislation.

### 3. Data Processor’s task

3.1. The Data Processor’s processing of personal data takes place in order to fulfil the Principal Agreement.
3.2. The purpose of the Data Processor’s processing is INSERT
3.3. The Data Processor’s task is to ...(e.g. host/support/operate the system) – clear description of the data processing carried out by the Data Processor, including how personal data is exchanged between the Data Controller and the Data Processor.
3.4. The Data Processor processes the following types of personal data:

e.g. name, address, personal registration number, health information (diagnoses, treatments etc.) This list must be as exhaustive as possible.

3.5. The Data Processor processes personal data regarding the following categories of data subjects:

e.g. employees from XX dept, patients with XX diagnosis, next of kin and test subjects,

as well as inclusion and exclusion criteria (when relevant).

3.6. The personal data covered by the Data Processing Agreement is processed (e.g. stored, hosted, backed up) at the following location(s), including ad hoc work premises:

INSERT

### 4. Technical and organisational precautions

4.1. The Data Processor must take all necessary measures required under article 32 of the General Data Protection Regulation. This article stipulates, among other things, taking into account the state of the art, the costs of implementation and the nature, scope, context and purposes of processing as well as the risk of varying likelihood and severity for the rights and freedoms of natural persons, the controller and the processor shall implement appropriate technical and organisational measures to ensure a level of security appropriate to the risk.

4.2. Appendix 1 (Data Processor’s instructions) stipulates the minimum requirements for the necessary technical and organisational security measures.
4.3. The principles and recommendations in ISO 27001, with later amendments, should be used as a framework guideline for fulfilling the Data Processing Agreement requirements.

### 5. Data Processor’s use of sub-processors

5.1. The Data Processor must not use another data processor (sub-processor) to process personal data covered by the Data Processing Agreement without the Data Controller’s prior written consent. Appendix 2 of the Data Processing Agreement lists the sub-processors approved by the Data Controller.
5.2. The Data Processor must notify the Data Controller and obtain specific written approval from the Data Controller if the Data Processor wishes to bring into use a new sub-processor. The notification must be received at least 3 months before the Data Processor wishes to bring into use the new sub-processor. The notification must take place by editing appendix 2 and sending it to.
5.3. In addition, at the request of the Data Controller, the Data Processor must provide a systemised overview of the entire chain of sub-processors mentioned in appendix 2, clearly indicating how the sub-processors are interrelated.
5.4. The Data Processor must enter into an agreement with a sub-processor which, as a minimum, complies with the data processing obligations undertaken by the Data Processor in the Data Processing Agreement. The sub-processor must have at least the same security level as undertaken by the Data Processor in the Data Processing Agreement.
5.5. If a sub-processor is used, the Data Processor must provide the relevant data processing agreement between the Data Processor and the sub-processor at the request of the Data Controller. The Data Processor must be able to document that the sub-processor has been instructed on the security requirements outlined in appendix 1 (Data Processor’s instructions).
5.6. The Data Processor is responsible for the contractual and legal requirements of the sub-processor’s processing of personal data. The fact that the Data Processor enters into an agreement with a sub-processor does not release the Data Processor from the obligation to comply with the Data Processing Agreement.
5.7. The Data Processor must inform the Data Controller of termination of an agreement with a sub-processor on processing of personal data covered by the Data Processing Agreement. In this case, the Data Processor must ensure that the sub-processor properly deletes all personal data in accordance with clause 10.
5.8. In the agreement with the sub-processor, the Data Processor must, when possible, include the Data Controller as preferred third party in the event of the Data Processor’s insolvency so that the Data Controller can assume the Data Processor’s rights regarding data processing and enforce them on the sub-processor. For example, so that the Data Controller can instruct the sub-processor to delete or hand back personal data.

### 6. Transfer of personal data to third countries or international organisations

6.1. The Data Processor must ensure that documented written approval is obtained from the Data Controller prior to the carrying out of transfers (e.g. assignment, disclosure or internal use) of personal data to third countries or international organisations unless the Data Processor is subject to other requirements under EU or Member State law. In such a case, the Data Processor must notify the Data Controller of this legal requirement before the transfer, unless the law prohibits this notification on important grounds of public interest. See General Data Protection Regulation article 28, section 3, paragraph a.
6.2. The limitations outlined in clause 6.1 mean, among other things, that the Data Processor must obtain prior written approval from the Data Controller to:
     i. Disclose personal data to a Data Controller in a third country or an international organisation.
     ii. Assign the processing of personal data to a sub-processor in a third country or an international organisation.
     iii. Allow personal data to be processed by another of the Data Processor’s departments located in a third country.
6.3. Providing that the Data Controller gives the Data Processor written consent to transfer personal data to a sub-processor in a third country, the Data Processor is responsible for ensuring that the personal data is not transferred unless there is a legal basis for transfer of personal data to the country in question. If a sub-processor in a third country is used, this must be stipulated in appendix 2 of the Data Processing Agreement.
6.4. If a transfer of personal data to a sub-processor in a third country, as outlined in clause 6.3, is based on the EU Commission’s Standard Contractual Clauses, the EU Commission’s Standard Contractual Clauses take precedence over the terms in the Data Processing Agreement for that specific transfer of personal data to the sub-processor. However, any requirements or obligations imposed on the Data Processor by the Data Processing Agreement and which are more onerous or which put the data subjects in a better position will still apply.
6.5. The Data Processor must, upon request from the Data Controller, provide any data transfer agreements entered into under the EU Commission’s Standard Contractual Clauses between the Data Processor and sub-processors.

### 7. Inspection and audit

The Data Processor must make available to the Data Controller all information necessary to demonstrate compliance with the General Data Protection Legislation and the Data Processing Agreement, and must allow for and contribute to audits and inspections carried out by the Data Controller or another auditor authorised by the Data Controller.

7.1. The following audits are in the Data Processing Agreement INSERT

7.2. The Data Processor is obliged to provide the supervisory authorities which, under applicable legislation, have access to the Data Controller’s and Data Processor’s facilities, or representatives acting on behalf of the supervisory authorities, with access to the Data Processor’s physical facilities on presentation of appropriate identification. The Data Processor is similarly obliged to ensure that this type of inspection can also take place at any sub-processors.
7.3. The Data Processor is obliged to inspect sub-processors. The Data Processor’s inspection of sub-processors must follow the agreed inspection procedure. The Data Processor must document on request that inspections of sub-processors have been carried out. Although the Data Processor is responsible for carrying out inspections of sub-processors, the Data Controller or a representative must have access to carry out inspections of sub-processors when necessary in the Data Controller’s professional opinion.

### 8. Notification obligation and assistance

8.1. The Data Processor is obliged to inform the Data Controller in writing and without unnecessary delay in case of any deviation from the Data Processing Agreement, for example:
     i. Any deviation from the instructions received
     ii. Any deviation from agreed access to personal data covered by the Data Processing Agreement.
     iii. Any deviation from planned releases, upgrades, tests etc. which are relevant for the processing of personal data covered by the Data Processing Agreement.
     iv. Any reasonable suspicion of confidentiality breach, misuse, loss or deterioration of personal data etc.
8.2. The Data Processor must inform the Data Controller without unnecessary delay when made aware of a personal data breach connected to the Data Processing Agreement at the Data Processor’s or at any sub-processors’ premises.

A notification of a personal data breach must include the following information:

i. The nature of the personal data breach and, if possible, who is affected, the number of affected data subjects, and the number of affected personal data records.
ii. Description of the likely consequences of the breach.
iii. Description of the measures the Data Processor has taken or suggests in order to manage the personal data breach, and what can be done to limit its possible adverse effects.
8.3. The Data Processor must assist the Data Controller as much as possible with the fulfilment of the Data Controller’s obligation to respond to requests for the exercise of the data subjects’ rights under chapter 3 of the General Data Protection Regulation, by providing appropriate technical and organisational assistance without unnecessary delay, and taking into account the nature of the processing.
8.4. The Data Processor must assist the Data Controller in fulfilling obligations for which the Data Controller is responsible under relevant legislation or when assistance is necessary in order for the Data Controller to fulfil these obligations, including the obligation to carry out a data protection impact assessment.

### 9. Agreement’s commencement and duration

9.1. The Data Processing Agreement enters into effect when signed by both parties.
9.2. The Data Processing Agreement is in force for as long as the processing of personal data is carried out and must remain in force until processing ceases.
9.3. The Data Processor and any sub-processors are obliged to hand back or delete personal data when data processing under the Principal Agreement ceases or at the written request of the Data Controller. The Data Controller must inform the Data Processor of the time at which data processing must cease. After this, it is the Data Processor’s responsibility to hand back or delete personal data at the request of the Data Controller, and also to delete existing copies at the reported time, unless EU or Member State law requires storage of the personal data.

### 10. Processing personal data after the agreement’s termination

10.1. The Data Processor is responsible for deleting personal data in such a way that it is not possible to recover the personal data. The Data Processor is responsible for deleting personal data from any backups and any sub-processors.
10.2. When data has been deleted, the Data Processor must send a written declaration that personal data has been deleted as agreed.
10.3. If the Data Processor or any sub-processors experience insolvency, liquidation or similar and therefore cease to process personal data for the Data Controller, all personal data must be immediately handed back to the Data Controller in a way which enables the Data Controller to continue making use of it. Following this, the Data Processor, the insolvent estate or similar is obliged to delete the personal data from own systems in accordance with the above.

### 11. Termination assistance

11.1. In the case of termination, cancellation or rescission of the Principal Agreement or the Data Processing Agreement, the Data Processor is obliged, at the request of the Data Controller, to provide termination assistance to the Data Controller until (i) all personal data has been transferred to the Data Controller in an ordinary, recognised electronic format, and (ii) the services described in the Principal Agreement have been satisfactorily handed over to the Data Controller or a new supplier selected by the Data Controller. The Data Processor must continue to process personal data and to deliver the services in accordance with the Principal Agreement until a satisfactory handover has taken place.
11.2. The Data Processor is obliged, at the request of the Data Controller, to inform the Data Controller in writing of how the Data Processor will transfer personal data and the services covered by the Principal Agreement to the Data Controller or a new supplier, including any technical requirements and conditions, so that the Data Controller or the new supplier can take over the services described in the Principal Agreement as well as the processing of personal data. The Data Processor must at the request of the Data Controller at any time be able to document and demonstrate to the Data Controller that the transfer of personal data and services can take place within the timeframe stated by the Data Processor.

### 12. Personal data covered by this agreement is confidential

12.1. The Data Processor must ensure and, at the request of the Data Controller, be able to demonstrate that all colleagues, sub-processors, business partners, external consultants and temporary workers etc. authorised to process the personal data covered by the Data Processing Agreement are bound by professional confidentiality or are subject to a relevant statutory duty of confidentiality.
12.2. The Data Processor is responsible for informing employees, sub-processors, business partners, external consultants and temporary workers etc. about the duty of confidentiality.
12.3. The Data Processor must ensure that access to personal data covered by the Data Processing Agreement is limited to employees who are required to process personal data in order for the Data Processor’s obligations to the Data Controller to be fulfilled. Access to personal data must therefore be blocked if authorisation is removed or expires.
12.4. The Data Processing Agreement’s obligations on confidentiality also apply after the Principal Agreement has terminated.

### 13. Transfer

13.1. The Data Processor must not transfer the rights and obligations of the Data Processing Agreement without the Data Controller’s prior consent.

### 14. Breach

14.1. In addition to the information in this clause (14), the Data Controller may invoke the usual remedies for breach provided by Danish law if the Data Processor breaches the Data Processing Agreement.

14.2. The Data Processing Agreement is part of the Principal Agreement, and therefore a breach of the Data Processing Agreement is also a breach of the Principal Agreement. In the case of material breach of the Data Processing Agreement, the Data Controller is entitled to cancel the Data Processing Agreement and the Principal Agreement.
14.3. The Data Controller’s cancellation of the Principal Agreement and the Data Processing Agreement does not mean that the Data Controller waives the right to compensation if the conditions for compensation are met.
14.4. Even if the Data Controller chooses not to cancel the Principal Agreement and the Data Processing Agreement on one or more occasions when entitled to do so, the Data Controller retains the right to cancel the Principal Agreement and the Data Processing Agreement on other occasions.
14.5. The Data Controller and the Data Processor are liable in accordance with the ordinary rules of Danish law in the event of breach of the Data Processing Agreement or violation of current General Data Protection Legislation.

If the Data Controller becomes liable to a third party, due to inter alia a claim of compensation, including a claim of compensation from the data subject(s) as a result of the Data Processor’s or any sub-processor’s breach of the Data Processing Agreement or violation of the current General Data Protection Legislation, the Data Processor must indemnify the Data Controller for all costs and losses. This is including cost and losses relating to injury caused to third party’s freedom, peace, honour or reputation.

Any liability or compensation limits determined in the Principal Agreement or elsewhere are not applicable in the case of the Data Processor’s breach of the Data Processing Agreement or violation of the current General Data Protection Legislation.

Article 82 of the General Data Protection Regulation is used to calculate compensation and other amounts payable to the data subjects as a result of a violation of General Data Protection Legislation.

The Data Processor’s obligation to indemnify the Data Controller in accordance with this section does not apply to fines or sanctions imposed on the Data Controller under article 83 or 84 of the General Data Protection Regulation.

14.6. The Data Controller is entitled to require the Data Processor to assist in protecting the Data Controller’s interests in any court or arbitration case, regardless of any objections the Data Processor may have about any alleged breach, as long as the Data Processor’s assistance is significant in the protection of the Data Controller’s interests, and as long as it does not harm the Data Processor’s position.

### 15. Applicable law and jurisdiction

15.1. The provisions in this section are not applicable if the applicable law and jurisdiction are regulated separately in the Principal Agreement.
15.2. The Data Processing Agreement and any questions about its validity are subject to Danish law.

15.3. Negotiation

If there is a dispute between the Parties concerning the Data Processing Agreement, the Parties must enter into negotiations with a positive, collaborative and responsible approach in order to resolve the dispute.

15.4. Court proceedings

If agreement cannot be reached by negotiation or otherwise, the dispute must be settled by the Danish court at the Data Controller’s domicile.

### 16. Changes to clauses 1 - 15

16.1. It has been necessary to make the following changes to the Data Processing Agreement [insert clause number]

### 17. Appendixes

Appendix 1: Data Processor’s instructions

Appendix 2: Sub-processors to the Data Processor

Any other appendixes

### 18. Signatures

On behalf of the Data Controller:

Name: Berit Schwartz

Job title: Head of Legal Department

Address: Blegdamsvej 9, 2100 København Ø

Date:

Signature:

The Data Controller’s project leader/system owner’s contact person:

Name: Carsten Utoft Niemann

Job title: MD

Date:

Signature:

On behalf of the Data Processor:

Name:

Job title:

Address:

Date:

Signature:

### Appendix 1 – Data Processor’s instructions

**Ref. 2. Data Processor’s responsibility**

2.2 Data processing covered by the Data Processing Agreement must take place in accordance with these instructions.

**Ref. 3. Data Processor’s task**

3.6 Ad hoc work premises
    3.6.1 2-factor authentication must be used. The authentication method can be, for example, Nem-id, SMS token, RFID or similar.
    3.6.2 External IT communication connections may only be established if agreed steps are taken to ensure that unauthorised persons cannot access personal data through these connections.
    3.6.3 The Data Processor must comply with the Data Controller’s guidelines for use of ad hoc work premises.

**Ref. 4. Technical and organisational precautions**

4.1 The Data Processor must, as a minimum, take the technical and organisational steps described below in connection with the processing of personal data covered by the Data Processing Agreement.
    4.1.1 If more detailed technical and organisational steps are necessary to ensure compliance with clause 4 of the Data Processing Agreement, these steps must always be taken.
4.2 Security risk
    4.2.1 The Data Processor must take the measures necessary to identify, evaluate and limit any reasonably foreseeable internal and external risks to the availability, confidentiality or integrity of all personal data covered by the Data Processing Agreement.

4.2.2 The Data Processor must take appropriate technical steps to limit the risk of any unauthorised access. The Data Processor must evaluate and improve the effectiveness of these precautions when necessary.
4.2.3 The Data Processor must document identified risks, as well as when a risk is reduced to an acceptable level.

The above obligation involves the Data Processor carrying out a risk evaluation followed by measures to counter identified risks. This could include any relevant measures from the following list:

a. Pseudonymisation and encryption of personal data
b. Capability to ensure continued confidentiality, integrity, availability and resilience of processing systems and services
c. Capability to correctly re-establish availability of and access to personal data in the case of a physical or technical incident
d. A procedure for regular trial, assessment and evaluation of the effectiveness of the technical and organisational measures for ensuring security of processing.
   4.2.4 The Data Processor must have formal procedures for handling security incidents.
4.3. Authorisation and access control
6. 4.3.1 Authorisations must state to which extent users may request, input or delete personal data.
   4.3.2 Only authorised persons may access personal data processed under the Data Processing Agreement.
   4.3.3 The Data Processor must be able to document which employees are authorised to access personal data processed under the Data Processing Agreement.
   4.3.4 Authorised persons must carry picture ID when processing data on-site at the Data Controller’s premises.
   4.3.5 Only persons engaged in purposes for which the personal data is being processed may be authorised. Individual users must not be authorised for uses they do not require.

4.3.6 Authorisation may also be given to persons who require access to personal data for auditing, operational or systems tasks.
4.3.7 Each authorised user is provided with a personal user ID and a personal password, which must be used every time the user accesses the data processing. Passwords must be changed every 6 months. Passwords must be sufficiently long and complex. Generally, 2-factor authentication must be used to access systems with sensitive personal data by internet or unsecured network. The authentication method can be, for example, Nem-id, SMS token, RFID or similar.
4.3.8 The Data Processor must take steps to ensure that authorised users can only access the specific personal data they have authorisation to access.
4.3.9 The Data Processor must have reasonable restrictions regarding physical access. Areas where personal data covered by the Principal Agreement is processed must be properly separated from general access areas.
4.3.10 The Data Processor must have formal procedures for dealing with resetting passwords, and other situations in which the normal, logical access controls are not in force.
4.3.11 There must be ongoing checks – at least once every 6 months – to ensure that users have the access and authorisation they should have. This check can include, for example, statistics created by the systems showing the individual users’ use of the systems so that it can be determined whether issued accesses and authorisations are still being used.
4.3.12 The Data Processor must, without unnecessary delay, cancel access and authorisation for users who, according to a concrete evaluation, no longer require them.
4.4. Training and instruction
2. 4.4.1 The Data Processor must make sure that colleagues receive adequate training and instruction to ensure that personal data is processed in accordance with relevant legislation as well as with the Data Processor’s policies and procedures.
4.5. Checks on failed access attempts and logging
4. 4.5.1 All failed access attempts must be registered. If 3 or more consecutive failed access attempts from the same user ID are registered within a set time, further attempts from that user ID must be blocked. Access must not be re-established until the reason for the failed attempts has been determined.
   4.5.2 All processing of personal data must be logged on hardware. The log must contain, as a minimum, information on the time, user, type of use, and identity of the data subject the data concerned, or the search criteria used. The log must be saved for 6 months and then it must be deleted unless the log’s purpose requires a longer storage period in order to use it as a tool in later investigation.
4.6. Input data materials
6. 4.6.1 Input data materials can only be used by persons who are engaged in inputting. Input data materials must be stored so that unauthorised persons cannot access the personal data within the materials.
   4.6.2 When it is no longer necessary to save input data materials, the Data Processor must delete or destroy the input data materials. The method for this must follow best practice.
   4.6.3 The provision regarding deletion or destruction does not apply if the material is covered by storage/discarding provisions related to other legislation, or if journalised materials are processed in accordance with the ordinary archiving provisions on storage, including the delivery of materials to the National Archives
4.7. Output data materials
8. 4.7.1 Output data materials are covered by the same instructions as input data materials, with the following addition:
   4.7.2 Output data may only be used by persons who are engaged in purposes for which the personal data is being processed, as well as for auditing, technical maintenance, operational monitoring and corrective measures etc.
4.8. Mobile storage media
10. 4.8.1 Mobile storage media with personal data must be clearly marked, and must be stored with an adequately strong encryption as well as under surveillance or locked up when not in use.

4.8.2 Mobile storage media with personal data may only be supplied to authorised persons for auditing or operational and systems tasks.
4.8.3 There must be a register of the mobile storage units used in connection with data processing.
4.8.4 There must be written instructions for use and storage of removable mobile storage media.

4.8.5 There must be sufficient necessary precautions taken in connection with repair and servicing of data equipment with personal data, as well as with the sale and discarding of used data media to ensure that personal data is not accidentally or deliberately destroyed, lost or deteriorated, and that personal data is not accessed by unauthorised persons, misused or otherwise processed in contravention of current legislation. This must happen following best practice.
4.9. Backups
2. 4.9.1 The same guidelines apply for backups as for all other processing of personal data described in this agreement.
   4.9.2 The Data Processor must ensure that systems and personal data are backed up regularly.
   4.9.3 Backups must be stored separately from the server in a non-adjacent room to ensure that they are not lost, for example in the case of fire or flood. Storage of backups must always be sufficient to avoid their loss.

4.9.4 The Data Processor must regularly check that backups are readable. This must include a perspective of preparedness – for example, for significant changes in a system’s technical set-up.
4.10. Updates and changes
2. 4.10.1 The Data Processor must have formal procedures to ensure that updates to operating systems, data bases, applications and other software are evaluated and implemented within a reasonable time.
   4.10.2 The Data Processor must have formal procedures for processing changes to ensure that every change is properly authorised, tested and approved before being implemented. The procedure must be supported by an effective separation of duties or management follow-up to ensure that no single person can implement a change alone.
4.11. Interruption in operations
4. 4.11.1 The Data Processor must have documented contingency procedures to ensure the re-establishment of services within a reasonable time in the case of an interruption in operations.
4.12. Disposal of equipment
6. 4.12.1 The Data Processor must have formal procedures in accordance with best practice as well as the Data Controller’s requirements to ensure that personal data is deleted effectively before disposal of electronic equipment.

4.12.2 When disposing of equipment, the Data Processor must document the methods used, and be able to produce this documentation upon request.
4.13 Inspection
     4.13.1 The Data Processor must carry out and document an inspection of the Data Processor’s organisation’s compliance with legal requirement, policies, procedures and this Data Processing Agreement with appendixes.

**Ref. 8. Notification obligation and assistance**

8.2	In the case of a breach of personal data security, the Data Controller must be informed in writing at the email address below without unnecessary delay so that the Data Controller can report the breach to The Danish Data Protection Agency and, if necessary, to the data subjects. The notification must be sent to:

The Capital Region of Denmark

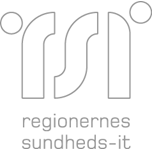


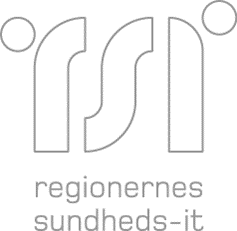

### Appendix 2 – Data Processor’s sub-processors

The Data Processor uses the following sup-processor(s) in connection with the tasks which the Data Processor carries out for the Data Controller. By entering into the Data Processing Agreement, the Data Controller approves the use of this sub-processor.

**One appendix per sub-processor.**

| **Sub-processor** |
| --- |
| Company’s full name |
| Company reg. number (or equivalent) |
| Company address (incl. country) |
| Other addresses which process personal data |
| Contact person at sub-processor |
| Does the Data Processor have an agreement with the sub-processor which fulfils the requirements of the Data Processing Agreement? |
| Data processing which the sub-processor participates in |
| Categories of personal data processed by the sub-processor |

| **Transfer of personal data to third countries** |  |
| --- | --- |
| Does the sub-processor process personal data in a third country? |  |
| If so, list the third countries |  |
| If so, provide the legal basis for the transfer (e.g. a standard EU contract or Binding Corporate Rules) |  |
| If the basis for the transfer is a standard EU contract, state which transfer model was used | ☐ Data Controller to Data Processor (with the Data Controller’s agreement)  ☐ Data Processor to Data Controller |
