## Appendix 2 for "The Danish Lymphoid Cancer Research (DALY-CARE) data resource: the basis for developing data-driven hematology"

**V. 4, 2nd OCT 2020:**
**Research Biobank and Clinical Database of patients with Lymphoproliferative Malignancies (LM).**

| | |
|---|---|
| WRITING COMMITTEE | Carsten U. Niemann, Caspar da Cunha-Bang, Michael Andersen, Christian Brieghel, Lone B. Pedersen, Rudi Agius, Torsten Holm Nielsen, Mette H. Thrane<br>Dept. Hematology 4042, Rigshospitalet<br>Copenhagen, Denmark |
| PROTOCOL SECRETARIAT & REGISTRATION | Dept. Hematology 4042 Finsen Center, Rigshospitalet<br>DK 2100 Copenhagen, Denmark<br>Att.: Carsten U. Niemann Fax (45) 35 45 48 41. Tlf: (45) 35 45 78 30<br> |

| | |
|---|---|
| TITLE | **A Research Biobank and Clinical Database of patients with Lymphoproliferative Malignancies (LM)**<br>A multi center study of LM patients followed from the time of diagnosis |
| PROTOCOL VERSION | 02 Feb 2020 |
| TRIAL TYPE | Research biobank and clinical database for patients with LM who have been diagnosed, entered a biobank, a research biobank or treatment biobank with LM in Denmark from 2002 till now and prospectively. |
| SUMMARY | |
| NO. OF PATIENTS AND SAMPLES | Presently: 700 from Rigshospitalet (CLL, jf sag 1405973, DNVK), further samples from biobanks at other hospitals will be requested, up to 2500 extra retrospective samples per year. Prospectively: 2500 patients annually. |
| INCLUSION AND EXCLUSION CRITERIA | All patients diagnosed with LM from 2002 with available samples stored in biobanks and/or available data. |
| TIMEFRAME | The project is expected to ongo until April 1st 2035 and thus samples will be stored until then |

<table>
<tr><td>OBJECTIVES</td><td>To study the integrated clinicopathological profile of LM: Anamnestic, clinical and paraclinical (biochemical, cellular, microbiologic, pathologic and genetic) profiles in relation to the potential course of treatment and outcome.</td></tr>
<tr><td>BACKGROUND</td><td>The present document represents an amendment to our previously approved protocol entitled “A research biobank of patients with chronic lymphocytic leukemia (CLL)” (1–3) (4) (5–7) (8,9) (10,11) (12) (13) (14). With this amendment, we wish to expand the study population of patients to include the entire spectrum of lymphoproliferative malignancies, not just CLL.<br><br>Lymphoproliferative malignancies (LM) cover a biologically and clinically heterogeneous group of diseases defined by the shared characteristic of aberrant proliferation of lymphocytes. For the purposes of this protocol, we consider only monoclonal/oligoclonal proliferation, not reactive/polyclonal lymphocyte expansion. Depending on the maturation stage at which the aberrant proliferation arises, different clinico-pathological entities can ensue. The spectrum of monoclonal LM, in its broadest sense, includes malignant diseases such as:<br><br>• Lymphoid neoplasms including: B-, T- and NK-cell lymphomas of all subtypes, CLL, multiple myeloma (MM) and related plasma-cell dyscrasias,<br>And,<br>• pre-malignant conditions such as monoclonal gammopathy of uncertain significance (MGUS) and monoclonal B-lymphocytosis (MBL) (15).<br><br>These diagnostic entities vary greatly in biology, clinical presentation, treatment strategy and, ultimately, outcome. However, common to them all is the background in the lymphoid immune system and thus deriving from the common cell type. We know from machine-learning  that integrating patient cohorts (16–18) and modelling them jointly, especially those with pre-existing commonalities, allows us to achieve predictive accuracies that are not achievable when modelling each LM separately. The current</td></tr>
</table>

| | |
|---|---|
| | amendment effectively extends the approach already successfully applied in CLL(19) to its neighboring diseases. Information to be investigated includes:<br><br>• Clinical information from patient records including information on treatment, outcome, infectious disease status as well as unrelated/secondary malignancies or transformation to other (typically more aggressive) LM diagnoses.<br>• Paraclinical information from blood samples, imaging, pathology review, microbiology, other laboratory analyses, cytogenetic analyses including next generation sequencing and other high throughput analyses.<br>• Information from functional research projects applying testing of biobank samples in assays such as mouse models, and in vitro assays.<br>• Microbiome analyses and other OMICs techniques.<br><br>Crucially, we plan to gather available information and integrate the analysis of diverse sources of information using machine learning as exemplified by our recent work in CLL (19,20). The integration of LM patient data will provide us with a unique platform for developing highly accurate prognostic models that are able to predict clinically relevant outcomes for patients with LM and also predict treatment responses.<br><br>(1,3) (2) (5–7) (8,9)(10,11) (12)(13)(14)The present protocol specifically aims at exploring anamnestic, clinical, molecular biology and paraclinical aspects in relation to the recorded outcome.<br>Such analyses may lead to new understanding of disease mechanisms, and aim for improved, individualized treatment. |
| PROJECT DESCRIPTION: TISSUE AND ANALYSES TO BE DONE | At Hematological Departments in Denmark all patients referred with suspected LM are offered routine diagnostic and prognostic work up which may include some or all of the following:<br>• Lymph node biopsy or other tissue biopsy |

| | |
|---|---|
| | • Imaging<br>• Bone marrow biopsy<br>• Blood samples<br>• Flow cytometry of blood or bone marrow aspirate<br>• FISH for certain well defined chromosomal aberrations<br>• NGS targeted sequencing<br>• Immunoglobulin heavy variable (IGHV) genes (part of the B-cell receptor): Somatic mutation analysis as compared to germline configuration. As part of this, IGHV-gene stereotypy has also been assessed.<br><br>In CLL, these studies have been done mostly on blood PBMNC (mononuclear cells mainly lymphocytes, sometimes also on bone marrow cells or biopsy tissue). The analyses are performed at the time of diagnosis and at times of disease recurrence/treatment need. Surplus PBMNC, bone marrow cells or biopsy tissue after analysis have subsequently been stored either as viable cells frozen in liquid nitrogen, as pellets of non-viable cells, as extracted DNA or parafin embeded as a treatment biobank. Based on the 1405973, DNVK approval of February 2015, patient samples have been collected for a research biobank upon informed consent from the patients while retrospective samples from the treatment biobank were transferred to the research biobank. This research biobank will form the research biobank for the current application for the Rigshospitalet site.<br>From 2015 and onward collection of other biological samples including saliva and fecal matter for microbiological analyses have also been performed on samples from CLL patients within the PERSIMUNE biobank for a subgroup of patients upon informed consent from the patients. With the current amendment, we wish to extend the collection of surplus samples of blood, bone marrow, tissue biopsy as well as saliva, skin swabs and feces to patients suffering from the entire spectrum of LM.<br><br>*The Danish Cancer Biobank* |

| | |
|---|---|
| | The Danish Cancer Biobank is a national cooperation between hospital departments handling tissue and blood samples from patients diagnosed with cancer.<br><br>*Local Treatment Biobanks*<br>Surplus material following biochemical, microbiological or pathological analyses are typically stored in local treatment biobanks in case reevaluation or additional analyses are needed later in the clinical course.<br><br>The materials include:<br>• Viable frozen PBMNC in liquid nitrogen at -180 º C<br>• Non-viable frozen PBMNC at -80 º C<br>• Non-viable frozen cell pellets at -20 º C<br>• DNA and RNA extracted for sequencing, frozen at -80 ºC.<br>• Plasma frozen at -20 ºC or -80 ºC.<br>• Biopsy tissue embedded in blocks of paraffin.<br>• Saliva, skin swabs and/or fecal matter at -20 ºC or -80 ºC.<br>The methodologies we wish to apply are:<br>1. BCR (IGHV) mutational status and stereotypy.<br>2. Fluorescence-in-situ hybridization (FISH), microarray and stimulated karyotype studies of specific gross genomic aberrations.<br>3. Somatic mutations of genes including Notch1, SF3B1, MYD88, TP53, ATM and other known recurrent somatic mutations by targeted next generation as well as Sanger sequencing and ddPCR.<br>4. Whole genome sequencing for detection of new coding and non-coding mutations and subclonal development in LM and sequencing for assessment of epigenetic regulation.<br>5. RNA sequencing for assessment of gene expression levels.<br>6. Flowcytometry including phosphoflow intracellular staining and BH3 profiling for pathway activation.<br>7. Mouse xenograft studies and in vitro assessment of primary CLL cells for prediction of treatment response and subclonal development upon specific treatment. |

| | |
|---|---|
| | 8. ELISA, western blotting and Multiplex bead array for cytokines and biomarkers in plasma and blodd/tissue.<br>9. 16S and whole metagenome shotgun DNA sequencing (mWGS) for assessment of microbiome.<br><br>Specifically, we wish to use both retrospectively and newly collected material from patients included at Rigshospitalet, at other hospitals or from clinical trials. Please see the table below for details on current molecular work up for patients with LM compared to the here proposed analyses.We also wish to use material from the project 1405973 which will be on going in this new project.<br><br>The number of already collected samples from clinical biobanks is aproximately 4000.<br><br>Samples will be stored until completion of the project which is expected at April 1st 2035.<br>Samples will be stored at the Department of Hematology, Rigshospitalet, Blegdamsvej 9, 2100. Section 4041. The samples will be stored safely behind 2 locked doors.<br><br>Methods:<br>1. IGHV mutational analysis: From a sample of anticoagulated blood we isolate DNA and/or RNA. The clonal IGHV rearrangement is amplified by PCR and Sanger sequenced. DNA sequences are analyzed in accordance with IMGT and IGBlast algorithms.<br>2. FISH and NGS analysis are performed by standard methodology at the Chromosome Laboratory, headed by Dr. Mette Klarskov.<br>3. Recurrent somatic gene mutations: By Sanger sequencing with published primers at the leukemia Laboratory gDNA or cDNA is analysed, selected exons PCR amplified and sequenced and compared with reference genes for mutations/polymorphisms. |

| | |
|---|---|
| | Assessment for recurrent mutations by ”Next Generation Sequencing” (NGS). The setup includes a design (Nimblegen) of a capture gene panel consisting of the relevant genes (incl. Notch1, SF3B1, Myd88, ATM, TP53, BIRC3 and other genes and regulatory elements reported with recurrent, somatic mutations in CLL) and for specific genes also of an amplicon-based assay. Briefly, DNA from patients will be fragmented, followed by insertion of index sequences and sequencing specific adaptors. The relevant genes will be extracted from the human genome and sequenced on the Illumina HiSeq 2500 platform or comparable platforms (at Genomisk Medicin). The data analysis will be done using the CLC Bio workbench and similar bioinformatics tools including custom designed algorithms for identification of variants/mutations.<br>4. For whole genome sequencing (identification of new somatic mutations in CLL), sequencing will be performed on Illumina HiSeq 2500 or similar platforms. Sequencing data will be processed using CASAVA-1.8.2. Trimming of the last 3’ base, mapping and variant calling will be performed by usage of CLC Genomics Workbench (Qiagen) and similar bioinformatic tools from which common variants (polymorphisms) are subtracted. Identification of disease causing/somatic mutations will be by Ingenuity Variant Analysis (Qiagen) and similar tools. Epigenetic features (including methylations) can also be analyzed by sequencing. By this approach, it is unlikely that germline mutations will be identified, as they will be called as disease related somatic mutations rather than germline mutations. However, if any information about inheritably mutations are identified by chance, this will be taken care of as described below in “ethical aspects”.<br>5. Flowcytometry including phosphoflow intracellular staining for pathway activation and BH3 profiling: Methods developed in the laboratory of A Wiestner, NIH, Bethesda, USA and the laboratory of A Letai, DFCI, Boston, USA.<br>6. Mouse xenograft studies: Model used and developed in the laboratory of A. Wiestner, NIH, Bethesda, USA, including work by CN (13). |

| | |
|---|---|
| | 7. Cytokines in plasma and biological samples, i.e. interleukines, measured by multiplex bead array and traditional ELISA techniques (21) and other proteomics approaches.<br>8. Microbiology analyses of saliva, biological samples and fecal matter using the 16S ribosomal RNA gene sequencing technique to identify and compare bacteria (16S) and a DNA sequencing method, whole metagenome shotgun (mWGS), that enables comprehensive sampling of all genes in all organisms in a microbial sample (22).<br><br>Clinicopathological correlates:<br>At diagnosis: Age, gender, performance status, hematology and biochemistry values (e.g. white blood cell count, lactate dehydrogenase (LDH), beta2-microglobulin (β2M), plasma immunoglobulin levels including free light chains), information on tumor burden (Lymph nodes as assessed clinically and on PET and CT scans, bone marrow and organ involvement, clinical stage, autoimmune manifestations and other complications. Some additional laboratory values (e.g. results of standard laboratory analyses like hemoglobin, platelet counts, creatinine, microbiology results, pathology reports etc) will be assembled through the PERSIMUNE datawarehouse set up according to our current data treatment agreement..<br>Follow-up data: Lymphocyte doubling time, time to first treatment, response to treatment, response duration, event-free survival, time to 2nd and later line treatment, respectively, overall survival, progression free survival, cause of death. Richter transformation. Severe infections, adverse events upon treatment and comorbidity. Other cancers. Autoimmune manifestations.<br><br>Collaboration:<br>As part of the project, some analyses will be performed by collaborators within Denmark and within the European Union as detailed below. These analyses will be part of the project described above. We apply for approval to exchange data and biologic material of the biobank with these researchers in order to build |

| | |
|---|---|
| | large enough patient series to detect rare correlations. For such an exchange, data will be pseudonymized, i.e. identified by a unique patient number, to which only the applicant and staff authorized by the applicant, has the key to translate into actual patient identity.<br>Rigshospitalet:<br>Next generation sequencing: Finn Cilius Nielsen, Genomic Unit, Rigshospitalet.<br>Denmark:<br>All hematological departments (Aalborg, Holstebro, Aarhus, Vejle, Esbjerg, Odense, Roskilde, Herlev and Rigshospitalet).<br>In the European Union:<br>Prof. Richard Rosenquist, Uppsala University, Sweden,<br>Prof. Anders Rosén, Linköping University Hospital<br>PhD Sigrid Skånland, Oslo University Hospital<br>Prof. Kostas Stamatopoulos, G. Papanicolaou Hospital, Thessaloniki, Greece<br>Prof. Paolo Ghia, San Raffaele Research Institute Milan, Italy,<br>Prof. Arnon P Kater, Akademisch Medisch Center Amsterdam, The Netherlands.<br>The international collaboration is anchored within the European Research Initiative on CLL (ERICLL.org) within which our laboratory is a reference center for TP53 mutations and certified for IGHV mutational analysis. For EU level data sharing based upon the ERICLL.org, we have an association with the HARMONY project on data sharing. The collaboration concerns projects covered within this research biobank with the need for more sample numbers to answer specific questions. Most projects concern sharing of specific data like outcome, baseline characteristics and specific recurrent mutations or IGHV sequences, thus not full exome sequences. For some projects with focus on specific mutations, sharing of biological samples like DNA or viably frozen cells will be included. For all projects, proper data treatment agreements including biobank samples if needed will be in place prior to project initiation. As an example of a joined EU project under way based on our prior research biobank, we have contributed targeted NGS sequencing data for a predefined list of 11 genes from 300 patient |

| | |
|---|---|
| | samples to a ERICLL.org based project on recurrent mutations in CLL, for which we use the data treatment agreement within the EU HARMONY project on big data in hematology.<br>Furthermore, the BH3 profiling may be performed at laboratory of M Davids, DFCI, Boston, USA based on a collaborative PhD project and a separate material transfer agreement. Any biological samples transferred to USA for this specific analysis will be further anonymized, i.e. a substitution of the study ID with a separate study ID only used for this part of the analysis. No other data on the patients from which such samples derive will be made available to the US laboratory.<br>In addition, metabolomic profiles may be performed through a collaboration with the lab of L Cerchietti, Cornell University, New York, USA based on a separate material transfer agreement. As above, any biological samples transferred to USA for this specific analysis will be further anonymized, i.e. a substitution of the study ID with a separate shudy ID only used for this part of the analysis. No other data on the patients from which such samples derive will be made available to the US laboratory.<br>For all collaborative ventures, data management agreements will be made. For collaborations with non-EU researchers, guarantees according to article #46 will be given.<br>The project has been approved by the data protection agency journal number 2012-58-0004, which will be updated to reflect the current amendment. |
| Statistical aspects | Univariate analyses of clinicopathological parameters will be assessed by Kaplan-Meier plots and log rank analyses.<br>Recurrent mutations: Associations between mutation rate and clinical features will be assessed by Wilcoxon rank-sum tests, Fishers's exact tests or Kruskall-Wallis tests. Multivariate analysis will be performed using General Estimation Equation and stepwise Cox proportional-hazards models. For call of disease specific mutations and gene expression analyses from RNA sequencing, statistical analyses will be performed as detailed for each technique above. For assessment of individual disease/treatment outcome, logistic regression and other complex mathematical models will be applied based on the above mentioned functional |

| | |
|---|---|
| | and genetic analyses. For pattern recognition Machine-learning methods like transfer-learning and multi-task learning will be applied (17,18). These algorithms are able to extract useful information from one disease to help predictions in another through the joint modelling of the lymphoproliferative malignancies. |
| OBTAINMENT OF INFORMED CONSENT | Retrospective part: For material stored in biobanks and research biobanks informed consent was collected at the time of sampling. For material collected as part of a treatment biobank, informed consent has not been obtained. Many of the patients included in the treatment biobanks over the years have died. We therefore apply for exemption from obtainment of informed consent from these patients.<br>Prospective part: Prospectively we wish to obtain informed consent from all new patients with LM at Rigshospitalet and other participating sites to donate blood and/or bone marrow, biological samples, saliva and fecal matter. Therefore, we apply for approval of the enclosed informed consent form.<br>As part of the informed consent (written and oral), the patient will be informed of the unlikely possibility of accidental information about possibly inheritable diseases and how such information will be disclosed to the patient, in the case the patient has indicated that (s)he wants to know about such information.<br>As part of the informed consent, the treating physician will review the study and the written informed consent together with the patient. The study will be explained in plain Danish language and the patient will have the opportunity to get response to all questions related to the study. There will be allocated due time for information of the patient in a calm and private environment as part of the outpatient consultation. The patient will be invited to have an accompanying person participating in the discussion of the informed consent and will be offered due time to discuss and review the information about the study before making a decision.<br>For patients with LM who are not any longer followed at a Department of Hematology in Denmark, they will be contacted by a letter (either electronically by eboks or by regular mail) informing about the research project (see attached example) along with the |

| | |
|---|---|
| | patient information. If the patient wants to participate or need further information, they will contact us to arrange an appointment. The patient will be able to give informed consent at the appointment or afterwards by letter. Hereafter an appointment for research samples will be made. For patients not responding to the first contact, a follow up letter with the same wording with an added reminder will be sent. |
| TRIAL SUBJECTS | Retrospective part: All consecutively newly diagnosed patients with LM with samples in biobanks, treatment or research biobanks from 2002 till now.<br>Prospective part: All consecutive, newly diagnosed patients with LM who, following informed consent, agree that their samples can be used for research.<br>Patients no longer followed in the outpatient clinic because of spontaneous regression can be contacted and included for follow up blood samples following informed consent. |
| RISKS AND BENEFITS | The research biobank does not per se imply any intervention, apart from potential extra samples of blood and – occasionally – bone marrow at time points where the patient will have these samples drawn anyway. There is only slight discomfort and no health risk associated with this. There is no direct benefit for the participants in the biobank. |
| SECURING OF PARTICIPANT DATA | The biobank has been approved by Datatilsynet under the Capital Region umbrella application (RH-2015-96 I-Suite nr.: 03856), and the demands of Datatilsynet concerning documentation of data security have been followed.<br>The laws on handling of personal information will be obeyed in accordance with the Danish Scientific Ethical Committees Act (Komitéloven) § 20, stk 1. nr. 4.<br>Patient samples will be coded with a unique registration number. Information about name and date of birth will only be found in a trial master file in a secure library (Rigshospitalets P-library).<br>Existing cases in the Danish Cancer Biobank will also be studied with available material.<br>Der vil indgå materiale fra det igangværende projekt 1405973<br>*nitrogentank CTS 2343 ARP, afsnit 4041, rum 4130, (aflåst)*<br>*afdeling: Hæmatologisk Klinik, Rigshospitalet, Blegdamsvej 9,* |

| | |
|---|---|
| | *2100 Kbh Ø)*, blod og knoglemarv. Tillige biologisk materiale i form af blod og knoglemarv samt mikrobiom samples (spyt og fæces) fra Persimune, Klinisk Immunologisk afdeling, afsnit 2032, Rigshospitalet. Herudover som anført biologisk materiale (blod, knoglemarv, lymfeknudevæv) fra biobanker på øvrige hospitaler.<br><br>Biological material from patients registered in the Danish ***Vævsanvendelsesregister*** will not be studied. |
| ETHICAL ASPECTS | The outcome of several types of LM has been strongly improved due to the increasing insight into LM biology and subsequent development of targeted therapy (23). Studies of tumor tissue correlated to the known course of disease of the participants can yield important new insights in disease mechanisms, which may further improve the outcome of LM and aid in the development of future treatment of the disease(s) (13,14).<br>The analysis of DNA mutations and RNA expression including mutations is based on tumor tissue. For all detected mutations that are expected to be inheritable, a group of experts from the department of clinical genetics and other relevant disciplines will be assembled. These experts with a broad knowledge of genetic diseases will then come up with a recommendation whether a specific patient should be informed about any chance findings. If so, genetic counselling will be offered to the patient.<br>In the case of accidental findings for patients within the treatment biobank, for whom an exemption from informed consent has been applied for, a likewise procedure will be implied. However, the expert panel will take into account for their recommendation that the patient has not had the opportunity to decide whether (s)he wants such information disclosed.<br>The use of xenograft mouse models and in vitro assessment of investigational agents on primary LM cells will open an important area of individually tailored treatment that can be tested in future clinical trials. Thus, the combination of molecular and cellular biology assays including NGS sequencing can be used to identify targets for new drugs that can subsequently be tested in primary cells and xenograft mouse models. Using this approach within our research group, that also performs phase I-III clinical trials, efficient |

| | |
|---|---|
| | translation of new insights from the laboratory into the clinic can be optimized – thereby benefitting patients the most.<br>The information gathered in this research project will solely be for scientific/statistical use. For prognostic models to be useful in a real-world clinical setting, highly accurate performance is necessary. Our previous work on CLL has shown that achieving high prediction accuracy is severely diminished when using a limited variety of sources to model patients (CLL-TIM REF). Naturally, we cannot expect to model the trajectory of a complex disease on a given patient by reducing them to a single snap-shot in time and a few variables. In fact, the reason why we have not seen major breakthroughs, in language translation, speech recognition and self-driving cars, translate into personalized treatment, is not due to lack of technology, but rather due to lack of integration of multiple data sources into predictive models. |
| RESPECT FOR THE STUDY PERSONS PHYSICAL AND MENTAL INTEGRITY AS WELL AS THEIR RIGHT TO PRIVACY | All samples procured to date serve the purpose of optimal management of this incurable disease, by preserving specimens that allow re-study at time of relapse and by comparison to new samples, identify new lesions and plan the optimal therapy. Clinical and paraclinical correlations have been done for some of the patients, participating in clinical trials duly approved by ethics committees and health authorities. We now apply for approval of this public research project for all patients in the biobank. Because the project includes genetic analyses of tumor cells and exemption from informed consent for patients from whom samples are obtained through treatment biobanks, this application is directed to the National Ethics Committee.<br>Biological material from patients registered in the Danish ***Vævsanvendelsesregister*** will not be studied. |
| Application for exemption from obtainment of informed consent | Local treatment biobanks have evolved as part of the routine patient management of LM, and not as result of a clinical trial warranting informed consent. Many of these patients have died. We therefore apply for exemption from obtainment of informed consent up to now regarding samples that are already in place. However, because we wish to continue to collect samples from existing patients and from future patients, we enclose a proposed patient information to be given to all existing patients from whom |

| | |
|---|---|
| | we wish to collect new samples and all new LM patients at diagnosis, and signed if informed consent is given. |
| ECONOMY | We will apply public and private foundations for the costs of the planned studies. We are currently applying for a large national grant (20 mio DKK) to secure the infrastructure to collect and assemble the biobank samples and data needed for the project, we have currently secured the following funding: |
| PUBLICATION | Both positive, negative and inconclusive research results will be published. Our group have a long track record of publishing these kind of data (19,20,24–29). |

| Post | Details | Amount | |
|---|---|---|---|
| Biobank logistics and PhD salary | Several private funds | 1580000 DKK | |
| Lab facilities and PhD salary | Kræftens Bekæmpelse | 1200000 DKK | Obtained from Knæk Cancer |
| Materials and postdoc salary | Novo Nordisk Foundation | 2500000 DKK | Obtained from clinical fellowship, 500000 /year 2017-2021 |
| Total | | 5280000 DKK | |

Table outlining current clinical work-up and research data point to be included for analysis in this proposal:

| Patient group | Current work-up | Expected results from WGS, standard | Expected results from WGS, research |
|---|---|---|---|
| **CLL** | IGVH rearrangements<br>Cytogenetic aberrations<br>TP53 mutations | IGVH rearrangements<br>Cytogenetic aberrations<br>TP53 mutations | Recurrent CLL mutations<br>B cell receptor (sub)clonality<br>MRD |

| | | | |
|---|---|---|---|
| **Multiple myeloma** | Cytogenetic aberrations identified by FISH (amp1q, del1p, del13q, del17p, 14q32 translocations t(4;14), t(11;14), t(14;16), t(14;20) | In supplementum to current FISH:<br>TP53 mutations (MM double hit)<br>BRAF mutations<br>MYC mutations | Recurrent MM gene mutations, including mutations in<br>- EGFR/MEK pathway<br>- Cereblon pathway<br>- NFkB pathway<br>- Epigenetic regulators<br><br>Molecular MRD monitoring |
| **B-NHL** | IgH gene rearrangements<br>Igϗ/Igλ light chain monoclonal expansion<br>Characteristic cytogenetic aberrations: t(11;14), t(14;18), MYD88, Bcl-2, Bcl-6, cMyc… | IgH gene rearrangements<br>Characteristic cytogenetic aberrations | ctDNA incl. MRD |
| **Hodgkin Lymphoma** | None | | ctDNA incl. MRD<br>Identification of novel disease-specific mutations and aberrations |
| **T lymphoma** | TCR rearrangements; patognomonic cytogenetic aberrations (e.g. t(2;5) in ALK+ALCL); specific rearrangements (e.g. DUSP22 in ALK-ALCL); immunohistochemistry | TCR rearrangements; patognomonic cytogenetic aberrations (e.g. t(2;5) in ALK+ALCL); specific rearrangements (e.g. DUSP22 in ALK-ALCL); mutations in TFH-derived T-cell | Identification of novel entity-specific mutations in genes not yet specifically associated with T-cell malignancies. Clonotypic TCR rearrangement and/or clonotypic mutational signature for ctDNA-based molecular MRD monitoring |

| | | |
|---|---|---|
| | (e.g. alk fusion protein) | lymphomas of epigenetic modifier genes (e.g. DNMT3A, TET2, IDH2) and RHOA |

19. Agius R, Brieghel C, Andersen MA, Pearson AT, Ledergerber B, Cozzi-Lepri A, et al. Machine learning can identify newly diagnosed patients with CLL at high risk of

infection. Nature Communications [Internet]. 2020 Dec [cited 2020 May 27];11(1). Available from: http://www.nature.com/articles/s41467-019-14225-8
